# Phenotypic and Genotypic Profile of Enterobacteriaceae Isolated at a Teaching Hospital in Ghana

**DOI:** 10.1101/2025.06.03.25328875

**Authors:** Bismark Donkor, Faustina Halm-Lai, Richael Odarkor Mills, Philimon Mwintige, Alberta Bedford Moses, Abigail Asmah Brown, Oheneba Charles Kofi Hagan

## Abstract

**Background:** Antibiotic resistance in *Enterobacteriaceae* continue to rise and its implications on health care delivery intensified. We investigated the genetic basis for antimicrobial resistance (AMR), virulence genes and associated plasmids in *Enterobacteriaceae* isolates at a teaching hospital in Ghana.

**Materials and methods:** Antimicrobial susceptibility testing was performed on archived isolates. Whole genome sequencing was performed on a subset of the isolates which were either multi-drug resistant or extend spectrum ß-lactamases (ESBL)producing. Bioinformatic analyses were performed for speciation, identification of AMR and virulence genes as well as associated plasmids.

**Results:** The 100 *Enterobacteriaceae* isolates included in this study expressed high phenotypic resistance to ß-lactams, and high susceptibility to aminoglycosides. The 20 WGS isolates were identified genotypically based on housekeeping genes *Escherichia coli* (8/20, 40%), *Klebsiella pneumoniae* (8/20, 40%), *Enterobacter cloacae* (2/20, 10%), and *Salmonella enterica* (1/20, 5%). These harboured 139 unique antibiotic resistant genes encoding resistance against ß-lactams (65/139), aminoglycosides (23/139), fluoroquinolones (45/139), tetracyclines (35/139), phenicols (28/139), and sulphonamides (9/139). Subsequent AST performed revealed that (74/79, 94%) were ESBL producers, and (9/79,11%) were CRE. The isolates expressed 8 main categories of virulence factors with adherence, effector delivery systems, and metabolic factors predominating in decreasing order. Additionally, 26 unique plasmid replicons of both I-complex and colicin plasmids were detected.

**Conclusion:** We identified marked phenotypic and genotypic evidence of antimicrobial resistance to commonly used antibiotics in the isolates at the hospital.

## Introduction

The *Enterobacteriaceae* group are widely distributed in nature, particularly in the environment and gastrointestinal tract of animal. They are Gram negative, non-spore forming, anaerobic, reduce nitrite to nitrate, and ferment glucose (1–3). Although majority of them are commensals, some genera have impacted on human and animal health. These include *Escherischia coli, Klebsiella spp, Citrobacter spp, Salmonella spp,* and *Enterobacter spp*(2,4,5).

The emergence and persistence of antimicrobial resistance (AMR) in the *Enterobateriaceae* to antimicrobials of various classes especially the extended-spectrum beta-lactams (ESBLs), carbapenems and fluoroquinolones in clinical settings have hampered global healthcare delivery especially in the lower- and middle-income countries (LMICs) 2020 (6,7). A systematic review and meta-analysis by Bezabih et al. found an eightfold increase in global antibiotic resistance enterobacteriaceae (ARE) rate, particularly, isolates that are ESBL carriage from 2003 to 2018 (8). Other observations present data that highlights the prevalence of CTM-X-15 gene, one type ESBL gene that is mostly dominant among pathogenic enterobacteriaceae (9,10). With regards to carbapenem resistance, the pooled prevalence globally is higher compared to data available in African. They report NDM, IMP and VIM-type as the main drive of CRE, especially NDM-1. However, data present in Ghana reveals low harborage of NDM-1 (11,12). Nonetheless, CRE in Ghana keeps on rising. Mills et al and Ayibieke et al reported that carbapenem-hydrolyzing class D β-lactamases, mainly OXA type genes are observed among enterobacteriaceae in Ghana (12,13).

Aside ARGs, these bacteria harbour genes of virulence factors (VFs). The VFs are important for the *Enterobacteriacea*e to successfully colonize and survive in their hosts (14,15), and eventually cause infections (7,16,17). VFs encompass diverse mechanisms such as host cell adhesion (eg. pili, adhesins), tissue invasion (eg. invasins, hyaluronidase), toxin production (eg. exotoxins, endotoxins), immune evasion (eg. capsules, antigenic variation), enzymes facilitating spread (eg. coagulase, proteases), iron acquisition (eg. siderophores), secretion systems (eg. Type III/VI), intracellular survival strategies, antiphagocytic (14,18,19) Surveillance of VFs among enterobacteriaceae reveals an abundance of VFs in ARE (14,18,20,21).

Both ARGs and VFs are harboured within bacteria chromosome and mobile elements such as plasmids, transposons and integrons (7,22–24). Present data reveals that they usually borne on plasmids (25,26), the dominant occurring mobile genetic element in antibiotic resistant bacteria (27,28). Incompatibility complex (I-complex) plasmids mostly consisting of resistance and fertility groups of plasmids are prevelance in ARE (25,29,30). IncF, IncC, IncX are reported to be the most I-complex plasmids involved in the transmission of AMR among Enterobacteriaceae (26,31,32). In addition to the I-complex plasmids, enterobacteriaceae also harbour colicin plasmids. These plasmids produce bacteriocins which have been reported to inhibit other bacteria in addition to being implicated in ARGs and VFs transmission (33).They are usually present in *Enterobacteriaceae*, especially *E. coli* strains and *Klebsiella spp* ((30,33,34).

Previous studies uncovered high record of *Enterobacteriacea*e infections in the CCTH (35,36); however, few data on the genetic basis of ARE, especially those that are ESBL producing is revealed at the hospital due to the poor antimicrobial resistance surveillance in Ghana. This study was undertaken to profile *Enterobacteriaceae* isolated from patients at CCTH. Through phenotypic identification and antimicrobial susceptibility testing (AST), we determined the prevalence of multidrug-resistant (MDR) strains, extended-spectrum β-lactamase (ESBL) producers, and carbapenem-resistant Enterobacteriaceae (CRE). Additionally, whole-genome sequencing (WGS) of 20 ESBL-producing isolates was performed to characterize their taxonomy, antimicrobial resistance genes (ARGs), virulence factors, and harboured plasmid replicons

## Materials and Methods

### Study design and site

This was a retrospective study of archived *Enterobacteriaceae* isolates from patients attending CCTH, a tertiary-level health facility located in the Cape Coast Metropolitan area of the Central Region of Ghana. The hospital is a 400-bed capacity facility which serves as a referral centre for the Central, Western, Western East and Eastern regions of Ghana. The microbiology laboratory at the hospital routinely undertakes culture and sensitivity testing for microorganisms isolated from various specimen sources including blood, urine, abscess, stool, wound swabs, high vaginal and endocervical swabs. The specimen was received from the various departments within the hospital including Enterobacteriaceae isolates archived from 2020 – 2023 were included in this study. the out-patient’s department (OPD), paediatric ward, the emergency wards, intensive care unit, and obstetrics and gynecology (O/G).

### Bacterial isolates

Bacteria isolates of the *Enterobactericeae* group were retrieved from storage and they were phenotypically identified using standard biochemical testing including citrate, oxidase, indole, urease and triple sugar iron (TSI).

### Antimicrobial susceptibility testing (AST)

AST was performed for the isolates using the following antimicrobial agents: (ß-lactams: ampicillin [AMP 10mcg], ceftriaxone [CTR, 30mcg], cefotaxime [CTX 30mcg], cefuroxime [CXM 30mcg], and meropenem [MRP, 10 mcg]; fluoroquinolones: ciprofloxacin [CIP 5mcg], levofloxacin [LEV 5mcg], and ofloxacin [OF 5mcg]; aminoglycosides: amikacin [AMK 30mcg], and gentamicin [GEN 10mcg]; sulphonamide: cotrimoxazole [COT 25mcg]; tetracycline [TET 30mcg]; and chloramphenicol [CHL 30mcg]. The AST was performed using the Kirby–Bauer disk diffusion method and the breakpoints were interpreted according to the Clinical and Laboratory Standard Institute (CLSI) 2020 guideline (37). Briefly, pure colonies of the cultured isolates were picked from agar culture plate and inoculated into peptone broth to achieve turbidity equivalent to 0.5 McFarland standards. Using a sterile cotton bud, a swab of the bacteria suspension was streaked over the entire surface of freshly prepared Mueller Hinton agar (MHA) plate. The antibiotic discs were placed on the MHA in the plates within 15 minutes of bacteria inoculation. The plates were incubated at 37 for 24 hours. Subsequently, the zone of inhibition was measured using a metre rule in millimeters, and interpreted as sensitive, intermediate or resistant according to the CLSI 2020 guideline (37). Based on the AST findings, we classified the isolates as either multidrug resistant (MDR)-resistance to 3 or more classes of antibiotics or ESBL producing.

### ESBL producers screening

Isolates exhibiting an inhibitory zone diameter ≤27 mm upon exposure to cefotaxime (30 mcg), and ≤25 mm when tested with ceftriaxone (CTR: 30 mcg) according to the (CLSI, 2020) guide were identified as possible ESBL-producers. ESBL confirmatory test was performed (n=79) using ceftazidime (CAZ:10mcg) and ceftazidime-clavulanic acid (CAL: 40mcg). Isolates that exhibited an inhibitory zone diameter ≤17 mm upon exposure to ceftazidime (30 mcg), and ≤20 mm when tested with ceftazidime-clavulanic acid were confirmed as ESBL producers.

### Whole genome sequencing (WGS)

Twenty isolates that were ESBL-producing were selected for WGS. Genomic DNA (gDNA) was extracted from the bacteria using the Quick-DNA mini prep plus kit (Zymo Research, Irvine, United States) according to the manufacturer’s instructions. WGS was performed at WACCBIP NGS Laboratory (Accra, Ghana) using the Illumina MiSeq (Illumina, Inc, San Diego, USA). DNA quality and quantity were determined using Qubit 4.0 fluorometer (Thermo Fisher Scientific, Waltham, USA). Short read sequencing was performed using (2 x 250 bp) PE sequencing with the MiSeq® Reagent Kit v3 (600 cycle). For each sample, 100 ng of total DNA was used for library preparation. Sequencing libraries were prepared using the Illumina DNA Prep (Illumina) library preparation kit from the enriched DNA following the manufacturer’s instructions. Using the Nextera XT Index Kit v2 (Illumina), distinct indices and Illumina sequencing adapters were attached to each individual library according to manufacturer’s instruction. Subsequently, each library was purified using Agencourt AMPure XP beads (BeckmanCoulter). The Agilent 4200 Tapestation (Agilent) was then used to check the expected size distribution and quality of the library. The concentration of the libraries was measured using the Qubit 4.0 fluorometer (Life Technologies). Based on the Tapestation and Qubit results, the barcoded libraries were normalized and pooled at an equimolar concentration. The combined library was diluted to 18 pM, spiked with 5% Phix (v3), and then sequenced.

### Bioinformatics analyses

Adapters and low-quality reads of (n=20) paired end (PE) reads were trimmed using Trimmomatic (0.39) (https://github.com/usadellab/Trimmomatic), setting Phred score at 33, ILLUMINACLIP: Nextera-PE. fa: 2:30:10, LEADING:3, TRAILING:3 and MINLEN:22. The PE trimmed reads (n=20) were assembled using SPAdes Genome Assembler (4.0.0) (https://github.com/ablab/spades), and quality assessment was assessed using QUAST (5.2.0) (https://github.com/ablab/quast). Multilocus sequence typing (MLST) (https://github.com/tseemann/mlst) which scans contigs against traditional PubMLST typing schemes, was performed to validate the identity of the isolates. Concurrently, each draft genome was screened for the presence of AMR genes using Abricate (https://github.com/tseemann/abricate) which host (CARD, PlasmidFinder, and VFDBS). Resistance and virulence genes were determined using CARD (38,39) and VFDBS (19) respectively. Assembled contigs were further assessed to identify plasmid genes using PlasmidFinder (40) in order to understand the transmission of AMR genes within the *Enterobacteriaceae* populace at the hospital. Genes were selected based on both coverage and identity equal to or greater than a threshold of 95%.

### Data Analysis

Data visualization was conducted using Python (v3.11) in Jupyter (v3.6.3) via Anaconda Navigator (v2.4.6), employing Matplotlib, pandas, and seaborn libraries. Heatmaps illustrating antibiotic susceptibility patterns and radar plots depicting virulence factor distributions among Enterobacteriaceae isolates were generated using the same libraries. Statistical analyses were performed in Jamovi (v2.3.28). Descriptive statistics summarized patient demographics and clinical characteristics, with medians and interquartile ranges calculated. Cohen’s Kappa was used to evaluate interrater reliability between phenotypic, MLST, and rMLST identification methods, implemented via the ClinicoPath module in Jamovi.

## Results

### Socio-demographics and clinical data

The majority of bacteria isolates were contributed by patients of whom (70%) were females, with 34% and 36% being within the age category of 20-29 and 40-69 respectively. The ages ranged from 8 days to 85 years with median age of 40 years and interquartile range of 35 years. Of the 100 samples 51% and 10% were specimen referred from the outpatient department (OPD) and accident and emergency (A&E) respectively. Other departments including IICU, FSW, MSW, and O/G contributed 13% of the isolates. Majority of the isolates (44%) were obtained from urine samples.

### Antimicrobial susceptibility patterns

Of the hundred (100) isolates, 6% were determined to be MDR. ESBL testing was successful for 79, out of which 74, that is, 93.7% were ESBL-producing. About 93%, 69% and 63% of the 100 isolates were susceptible to amikacin, chloramphenicol and gentamicin, however, all the isolates were resistant to ampicillin (**Fig1**). Meropenem sensitivity testing was successful for 79 of the isolates out of which 40.51% (32/79), 48.10% (38/79), 11.39% (9/79) were either susceptible, intermediate or resistant respectively: This suggests that the isolates are mostly susceptible to meropenem at the hospital.

**Fig 1:**
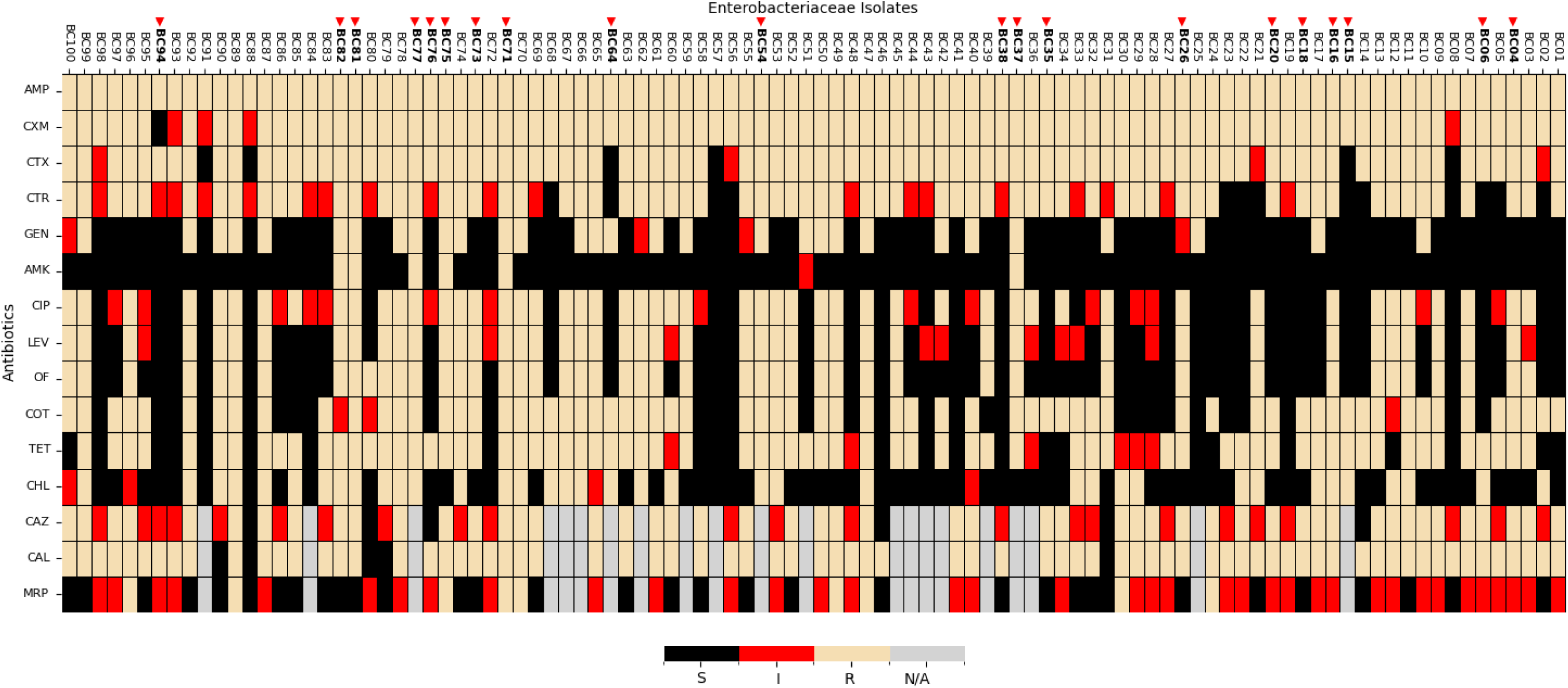
Antibiotic susceptibility profiles (phenotype) of *Enterobacteriaceae* bacteria Isolates. The heatmap displays antibiotic susceptibility patterns across various *Enterobacteriaceae* isolates for different antibiotics. Each row represents a specific antibiotic, while each column corresponds to an isolate. The color-coding indicates the susceptibility categories: black: susceptible (S), red: intermediate resistance (I), beige: resistant (R), gray: data not available (N/A). The antibiotics tested were (ß-lactams: Ampicillin [Amp], Ceftriaxone [CTR], Cefotaxime [CTX], Ceftazidime [CAZ], Meropenem [MRP], and Cefuroxime [CXM]; Fluoroquinolones: Ciprofloxacin [CIP, Levofloxacin [LEV], and Ofloxacin [OF]; Aminoglycosides: Amikacin [AMK], and Gentamicin [GEN]; Sulphonamides: Cotrimoxazole [COT]; Tetracycline [TET]; Chloramphenicol [CHL]; and Ceftazidime-Clavulanic acid [CAL]).

### Whole genome assembly

Computation of the total read count and quality metrics of the assemblies (**Additional File 4**) were consistent and of high-quality. Tseeman/mlst analysis predicted *E. coli* (8/20, 40%), *K. pneumoniae* (8/20, 40%), *E. cloacae* (2/20, 10%), and *S. enterica* (1/20, 5%). Additionally, (1/20, 5%) was identified as *Pseudomonas aeruginosa* which belongs to the family *Pseudomonadaceae* (**Additional file 1**). Agreement between the phenotypic and genotypic identification of the *Enterobacteriaceae* isolates, and Kappa were found to be moderate with 0.60 and 0.47 respectively. The statistical difference between the phenotypic and genotypic identification of the *Enterobacteriaceae* isolates was significant (p-value < .001). It suggests strong evidence that the phenotypic and genotypic methods, including MLST and rMLST, do not consistently agree in their classification of bacterial isolates.

### Resistome

CARD revealed 139 unique ARGs that would confer resistance to the following classes of antibiotics: ß-lactams (65/139), fluoroquinolones (45/139), aminoglycosides (23/139), phenicol (28/139), tetracycline (35/139), and sulphonamide (9/139) as shown in table 2. High diversity of ARGs were observed in *E. coli* (130/139) followed by *K. pneumoniae* (111/139), *E. cloacae* (9/139), and *S. enterica* (6/139), respectively. Isolate IDs BC82 (*E. coli*) harboured most ARGs (82/139) whereas lowest was detected in both BC76 and BC94 (*K. pneumoniae)* (4/139) each. *CTX-M-15*, an ESBL gene was the most observed ARG, detected among (13/19) isolates. Nine ARGs including *APH (3’)-Ia, Erm* (*49*)*, EC-15, Escherichia_coli_emrE, golS, mdsC, MdtK, QnrD1,* and *SHV-80* were recorded as the lowest occurring ARGs among the isolates. Each were observed once in just a single isolate. Several variants of multidrug resistant genes (91/139) were observed in the isolates, especially in *E. coli* (65/91), followed by *K. pneumoniae* (44/91), *S. enterica* (5/91), and *E. cloacae* (1/91), respectively. They were observed to have conferred resistance, especially on 4 classes of antibiotics including ß-lactams, fluoroquinolones, tetracycline, and chloramphenicol.

**Table 1:** Demographics and clinical characteristics of isolates corresponding to patients at CCTH.

| Category | Sub-category | Percentage (%) |
| --- | --- | --- |
| **n =100** |  |  |
| **Age in years** | ≤12 years | 12.0 |
|  | 13-19 | 2.0 |
|  | 20 -39 | 34.0 |
|  | 40- 69 | 37.0 |
|  | ≥70 | 15.0 |
| **Sex** | Female | 70.0 |
|  | Male | 30.0 |
| **Department** | OPD | 51.0 |
|  | Other Departments | 13.0 |
|  | A/E | 10.0 |
|  | FMW | 8.0 |
|  | MMW | 7.0 |
|  | ETAT | 6.0 |
|  | PMW | 5.0 |
| **Specimen Source** | Urine | 44.0 |
|  | Sputum | 14.0 |
|  | HSV | 12.0 |
|  | Wound Swab | 12.0 |
|  | Other Sources | 10.0 |
|  | Blood | 8.0 |

OPD: Outpatient Department; A/E: Accident & Emergency; FMW: Female Medical Ward; MMW: Male Medical Ward; ETAT: Emergency Triage, Assessment & Treatment; PMW: Paediatric Medical Ward.

**Table 2:**
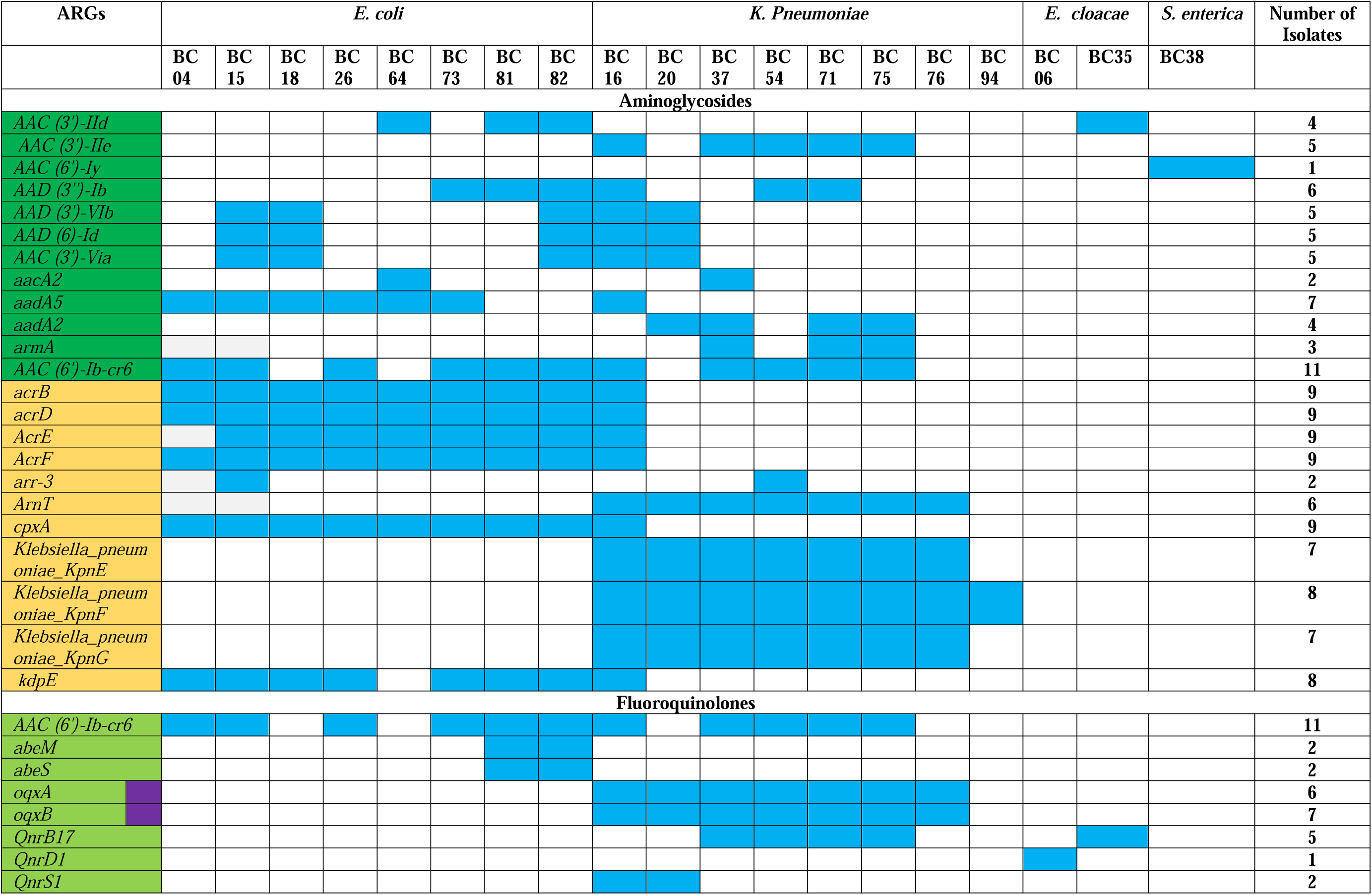
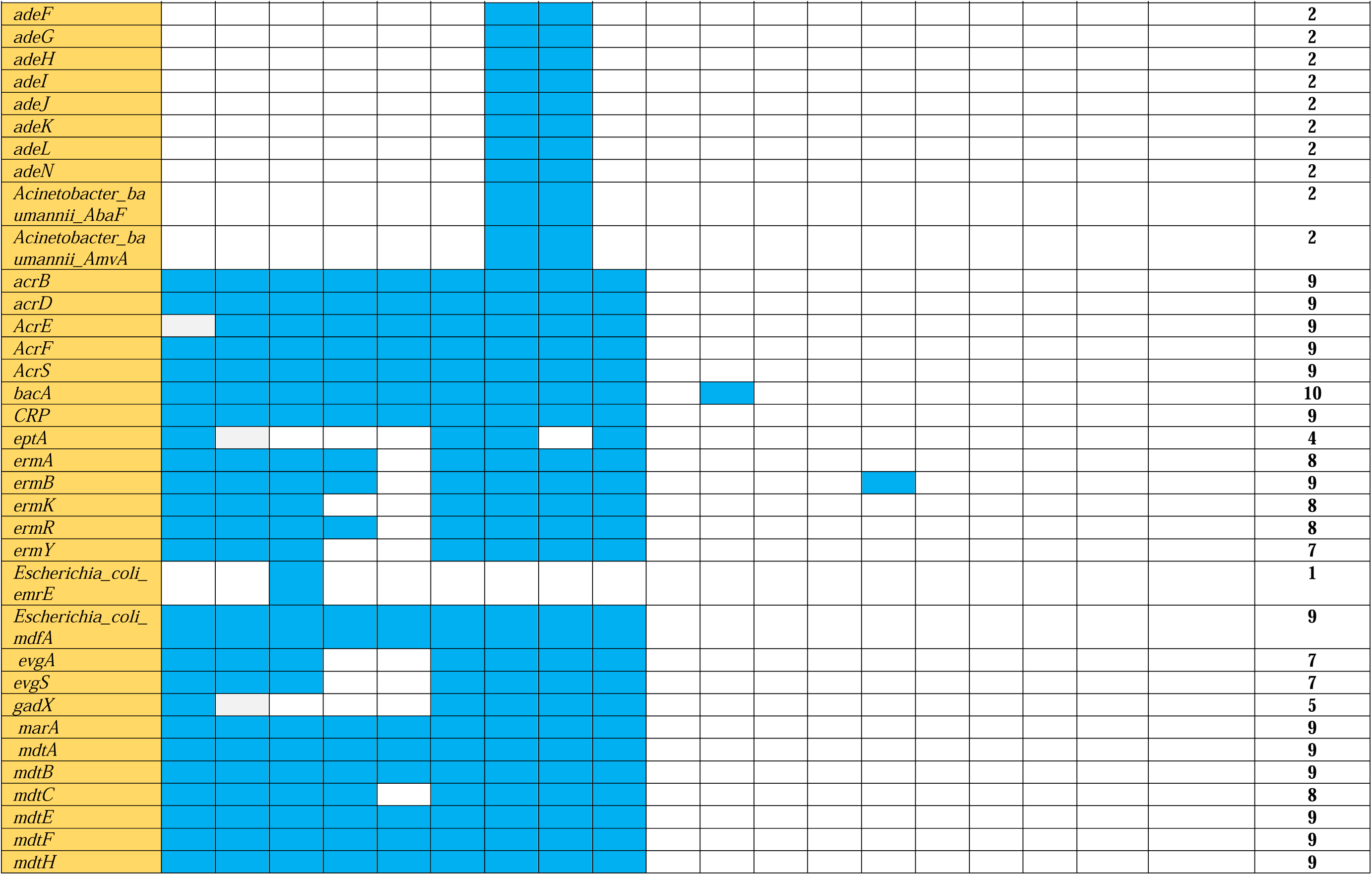
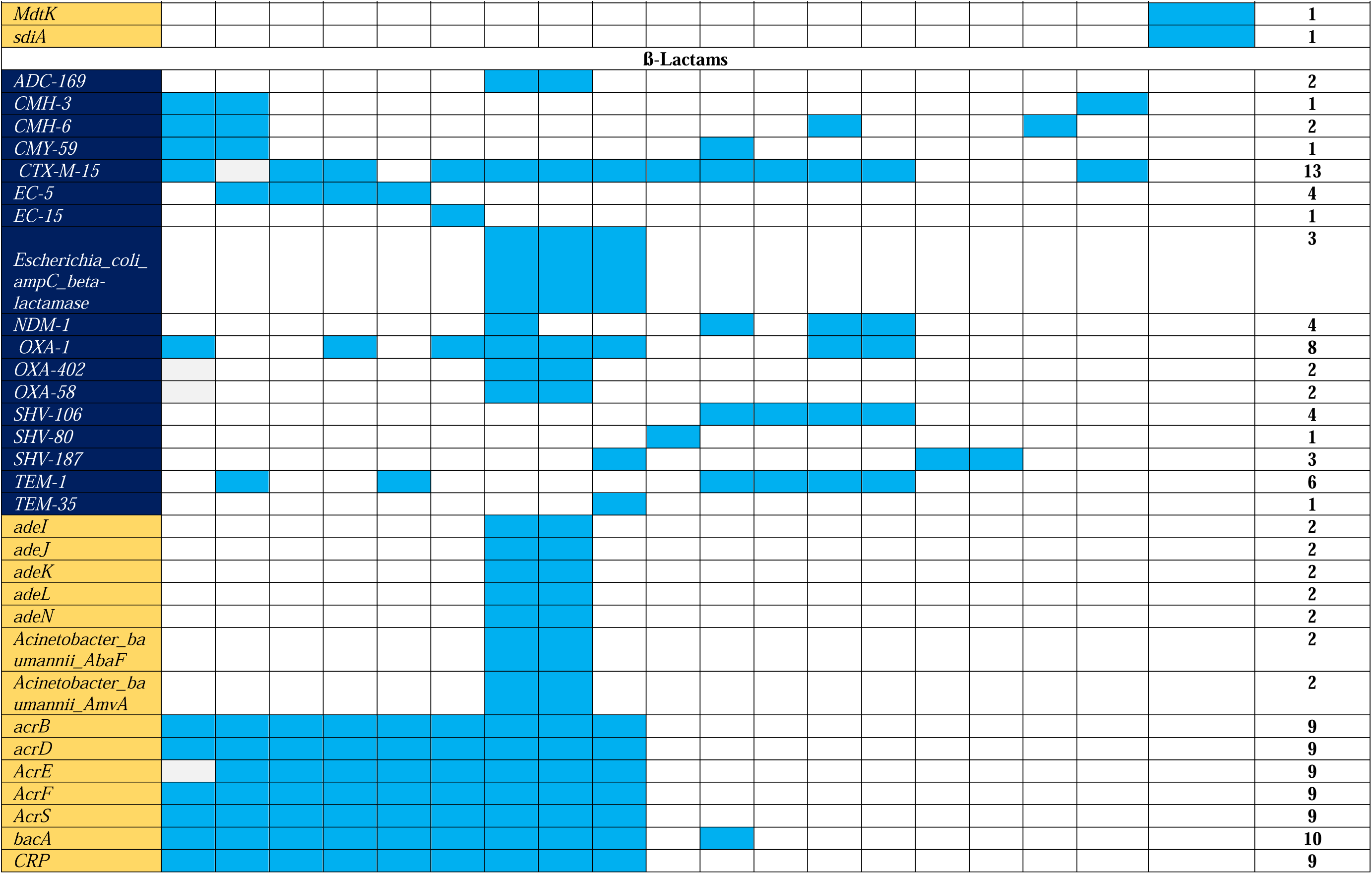
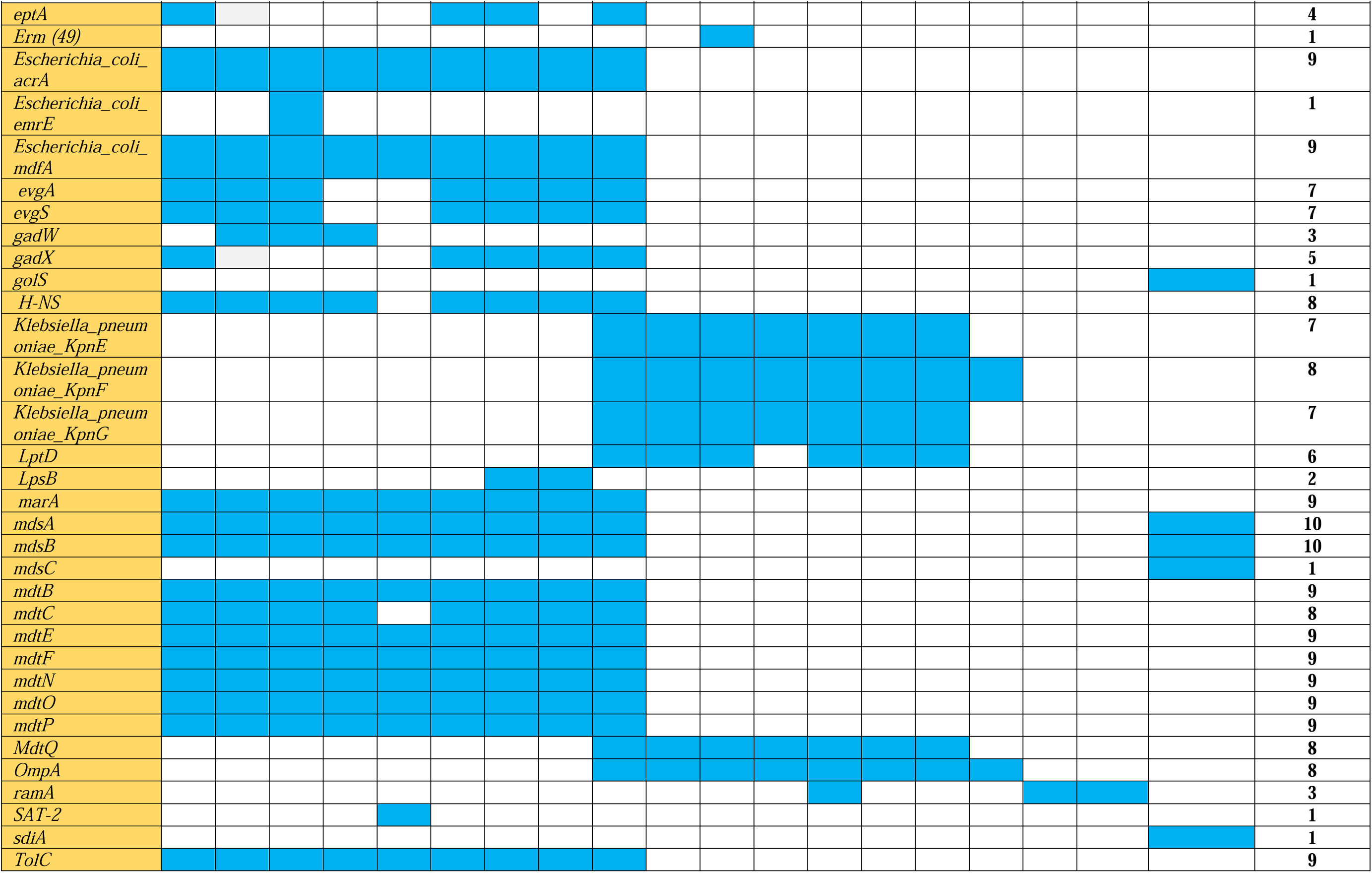
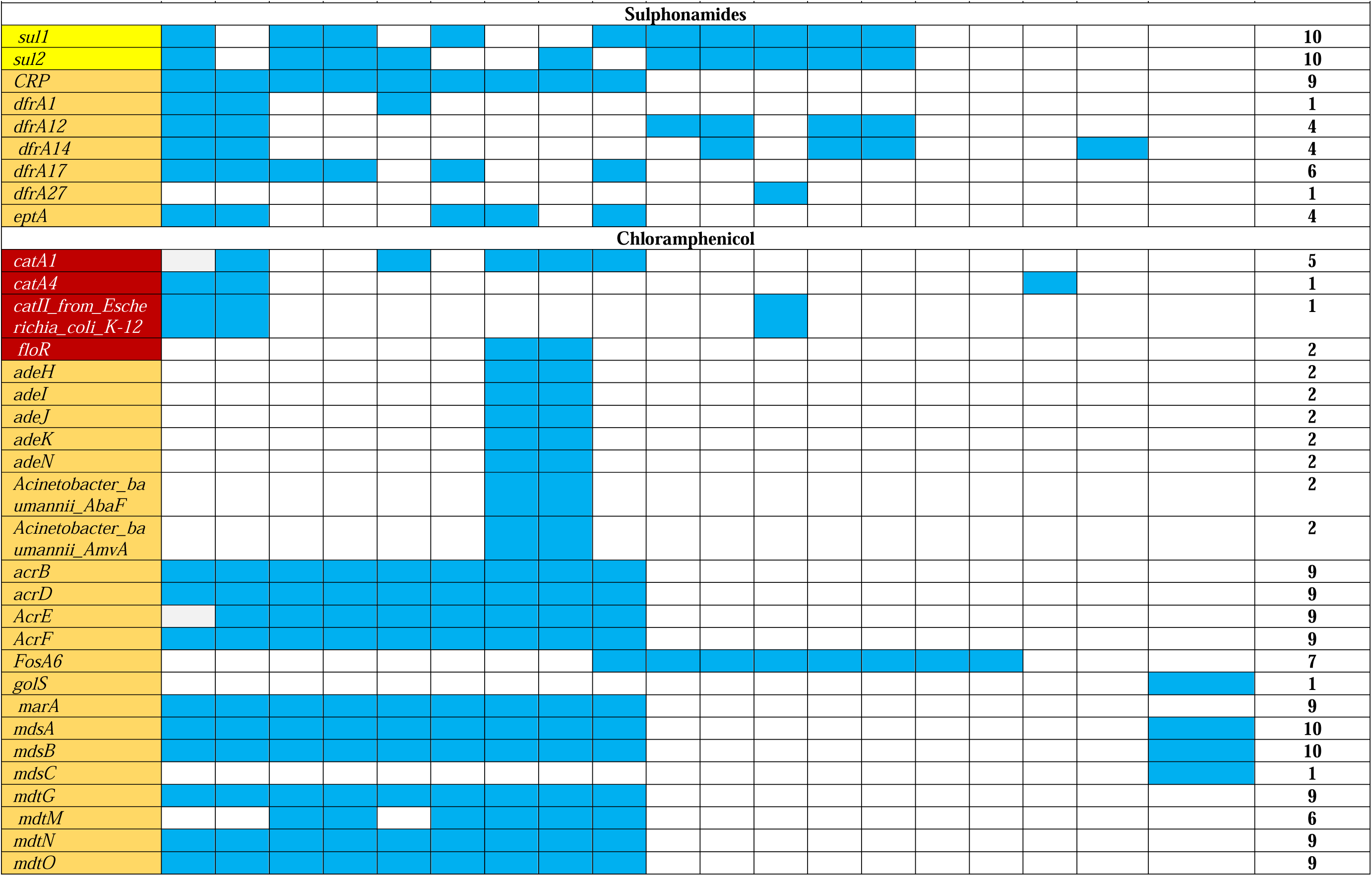
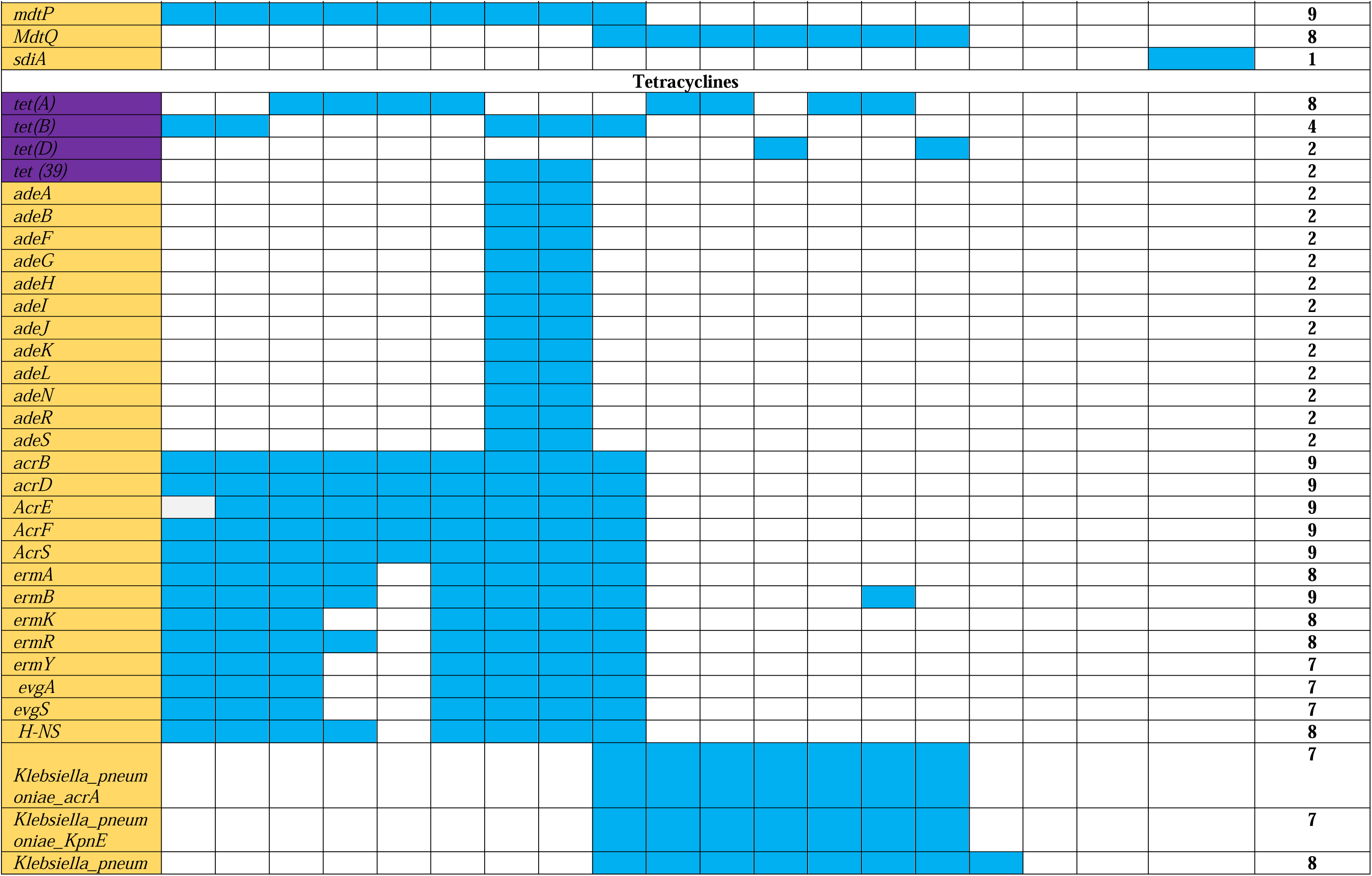
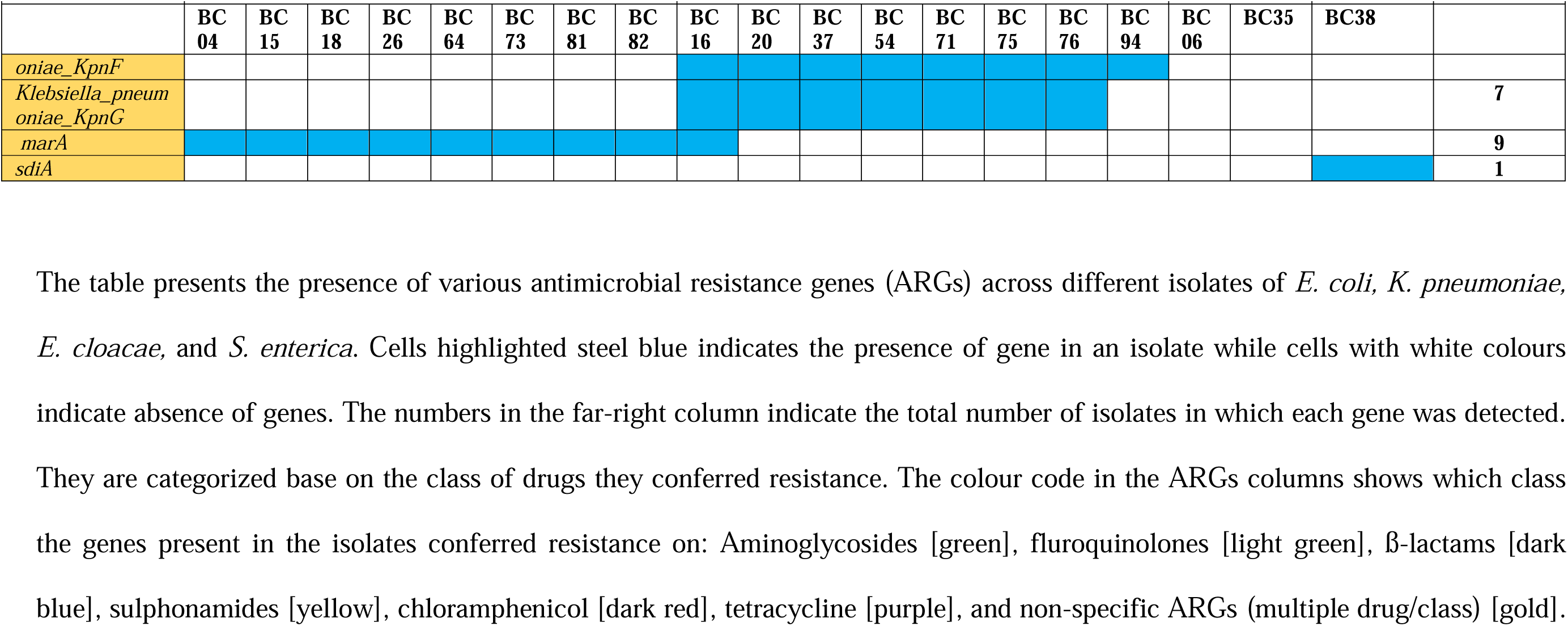
Presence of antimicrobial resistance genes (ARGs) in *Enterobacteriaceae* bacterial isolates.

### Virulome

Using the VFDBS several VFs previously described in the *Enterobacteriaceae* family (*E. coli, K. pneumoniae, E. cloacae, S. enterica*) were detected. In total, 414 unique virulence genes were detected (Additional file 3), categorized into 8 major groups and 17 subcategories, as detailed in Table 3 and Fig 2. The predominant VF categories included adhesins, invasins, effector delivery systems, exotoxins, immune evasins, nutrient and metabolic factors, biofilms, and exoenzymes. Majority of these VFs were observed in E. *coli* isolates, followed by *K. pneumoniae, E. cloacae,* and *S. enterica.* Adhesin-associated VFs, essential for bacterial attachment to host cell mucosae (41), were predominantly identified in *E. coli* and *K. pneumoniae*. Invasion-related VFs, which facilitate host cell penetration, were detected across all four genera. Effector delivery systems, including T2SS and T3SS, known for injecting bacterial proteins into host cells to manipulate immune signaling, cell death, and nutrient acquisition pathways (42,43) were also observed in all the genera principally in the *S. enterica* isolate as shown in *Fig 2*. Exotoxins particularly membrane-acting, intracellular active toxins were observed in *E. coli, K. pneumoniae*, and *E. cloacae* isolates, however, none was observed in the *S. enterica isolate*. Similarly, immune evasion VFs, including antiphagocytosis and complement evasion mechanisms, were present in all Enterobacteriaceae isolates except *E. cloacae.* Nutritional and metabolic VFs were present mostly in *E. coli* followed by *K. pneumoniae and E. cloacae*. These factors, particularly those involved in metal uptake, metabolic adaptation, and iron sequestration, are crucial for bacterial survival and proliferation in host environments (20,44). Biofilm-associated VFs, which contribute to antibiotic resistance, were confined to *E. coli and K. pneumoniae*. Meanwhile, proteases that degrade exogenous proteins, enhancing bacterial growth and metabolism (45), were uniquely observed in *E. cloacae*. Bacterial survival in host environments plays an important role in persistent infections (46). *Enterobacteriaceae* strains harbouring these VFs can subvert host immune defenses, leading to recurrent infections (14,47). The diversity of VFs identified in the *Enterobacteriaceae* isolates accentuate heir adaptability within host environments, enhancing their ability to evade antimicrobial defenses and establish infections.

**Fig 2.**
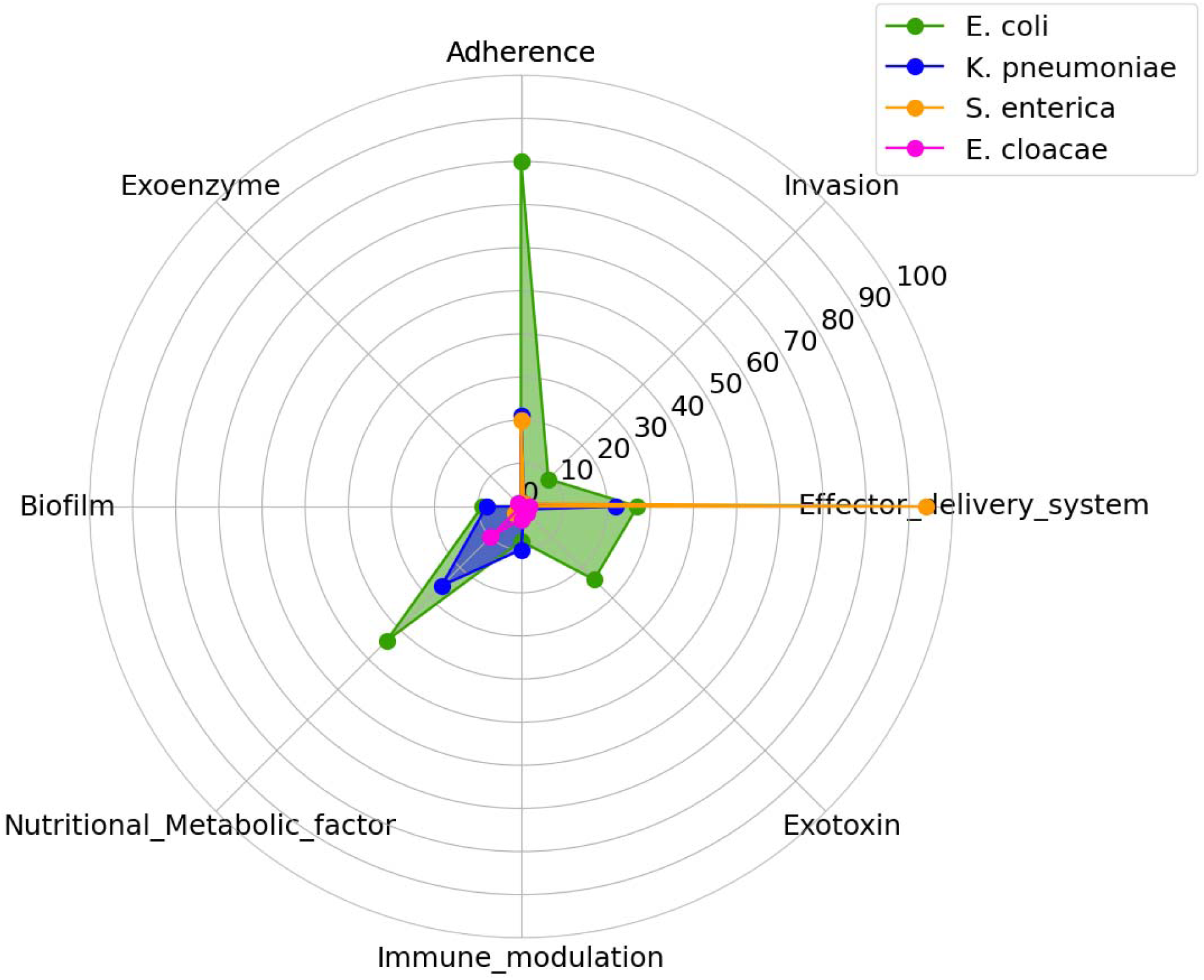
Virulence factor distribution among *Enterobacteriaceae* bacterial species. This radar chart illustrates the distribution of eight virulence factors (Adherence, Invasion, Effector delivery system, Exotoxin, Immune modulation, Nutritional/Metabolic factor, Biofilm, Exoenzyme) across four *Enterobacteriaceae* bacterial species: *E. coli* (green), *K. pneumoniae* (blue), *S. enterica* (orange), and *E. cloacae* (pink). Each axis represents a virulence factor, with the distance from the center showing its strength or pathogenic and AMR contribution in each species. *E. coli* stands out in Adherence and Nutritional/Metabolic factors, while *S. enterica* is strong in the Effector delivery system. The other species show varying levels of these factors, reflecting their different pathogenic profiles.

**Table 3:**
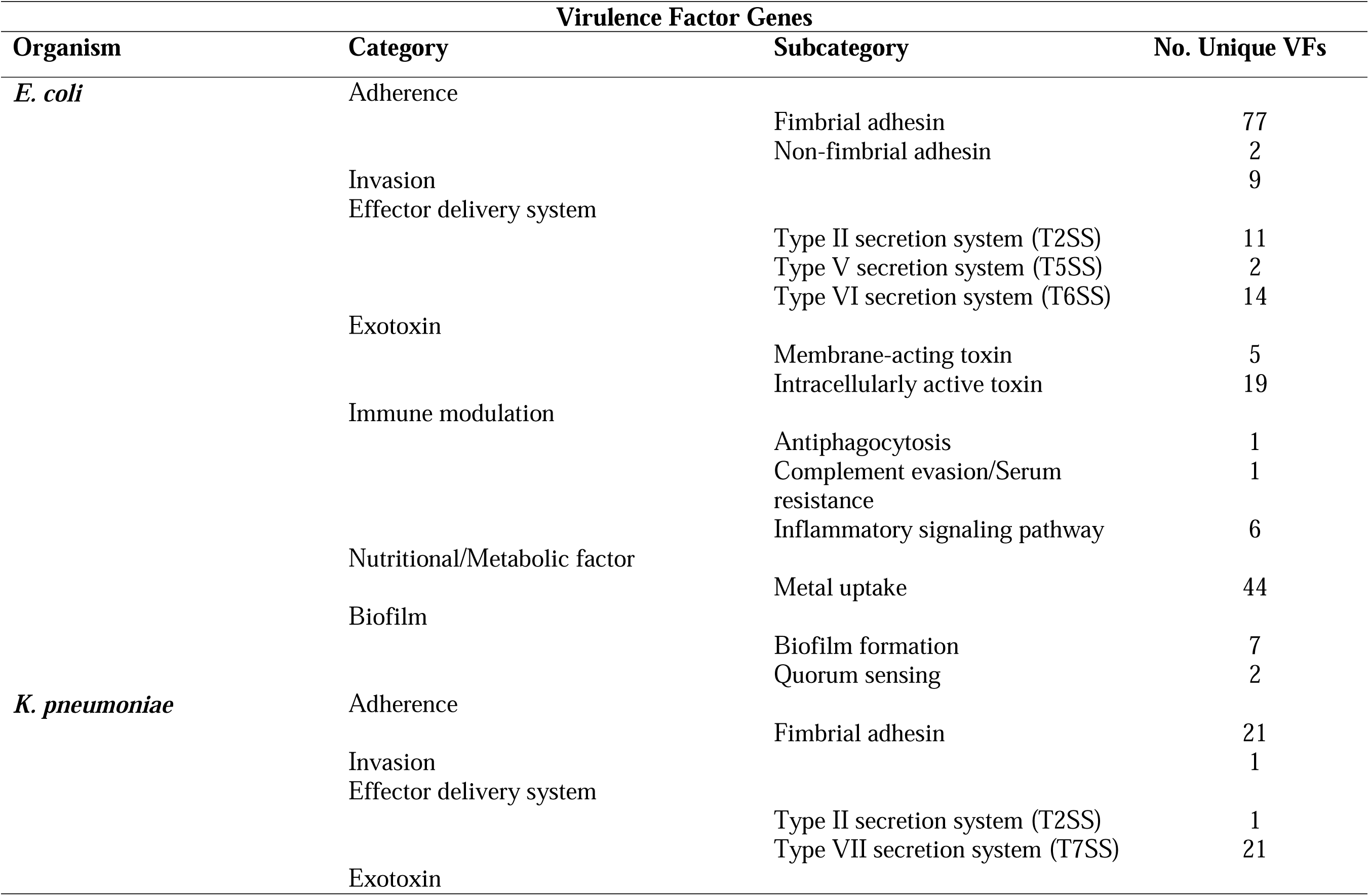
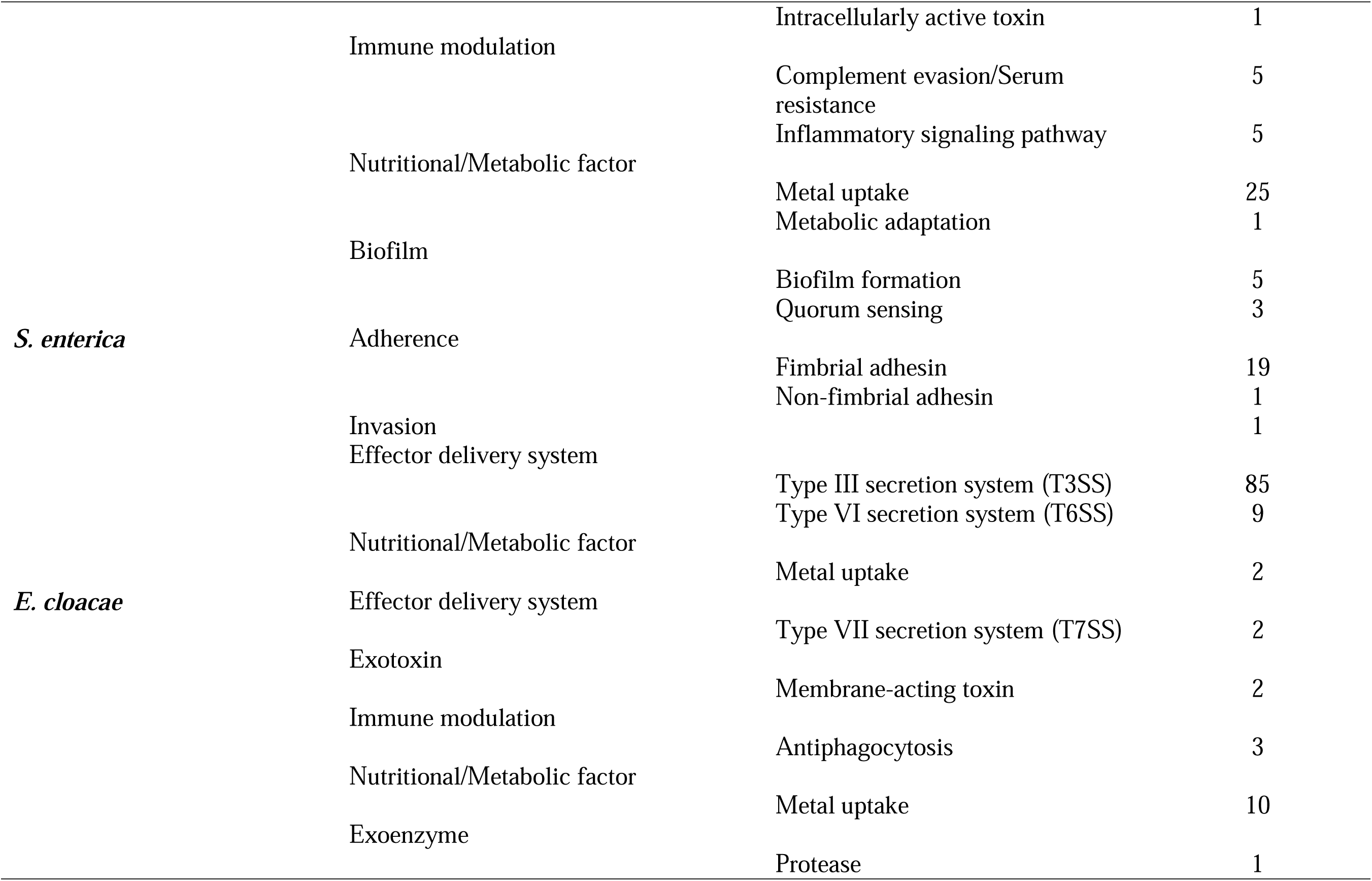
Virulence factors and sub categories present among Enterobacteriaceae bacterial isolates.

### Plasmidome

The results from Plasmid Finder revealed a diverse array of plasmid types classified under Incompatibility (Inc) complex plasmids and Colicin (Col) plasmids as shown (table 4). The number of plasmid replicons per isolate varied significantly. *K. pneumoniae* exhibited the highest diversity of plasmids (18/26) followed by *E. coli* (9/26), and *E. cloacae* (5/26). The highest plasmid diversity was observed in isolate BC37 of *K. pneumoniae* (8/26), whereas BC82 (*E. coli*) and BC26 (*K. pneumoniae*) exhibited the lowest plasmid diversity (1/26 each). The only *S. enterica* in this collection harboured no plasmid replicon. Among the Inc plasmids which are mostly observed in *Enterobacteriaceae* as vehicles for AMR transmission (29), IncF subgroups were the most frequently detected. *IncFIB (AP001918) _1* was the most prevalent, identified in 8 isolates, followed by *IncFIB(K)_1* and *IncFIA_1*, which were found in 7 isolates each. *IncFII_1, IncHI1B(pNDM-MAR) _1*, and *IncR_1* was detected in 3 isolates each. Plasmids such as *IncC_1, IncFII* (*29*) *_1*, *IncFII(pCoo)_1, IncX4_2,* and *pKPC-CAV1321_1* were found in only a single isolate each. Colicin plasmids which encode colicins (bacteriocins), proteins that kill other bacteria (30) were also distributed across the isolates. They are reported to contribute to the spread of antibiotic resistance and other virulence factors among uropathogenic *E. coli* strains and *Klebsiella spp* by co-integrating with other plasmids like *IncC, IncF, and IncN*, (33,48). *Col156_1* was the most frequently detected (4). *ColRNAI_1* was present in 3 isolates, while *Col (BS512) _1 and Col440I_1* was each observed in 2 isolates. The least common Colicin plasmids, found in single isolates, included *Col (KPHS6) _1, Col3M_1,* and *ColpVC_1*.

**Table 4:**
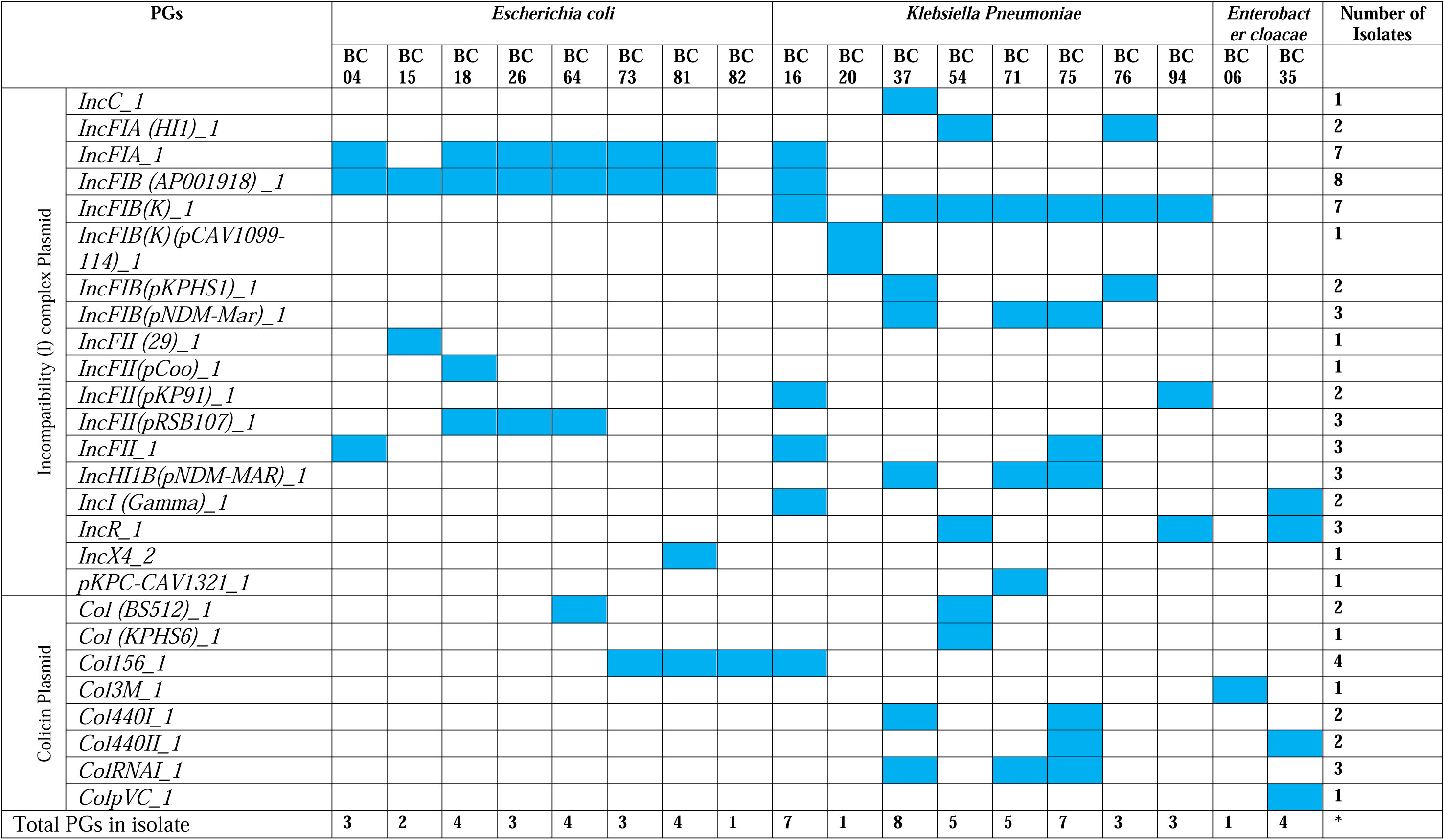
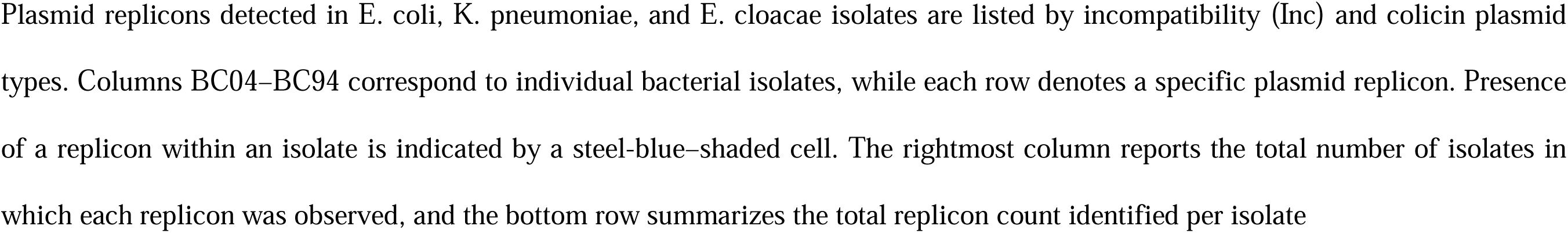
Plasmid replicons present among Enterobacteriaceae isolates.

## Discussion

Phenotypic surveillance of bacterial infection in the clinical settings complemented with genomic studies enable the elucidation of basis of diseases cause, accurate identification, transmission dynamics and AMR (49,50). Our study has highlighted the need for the adaptation of genetic surveillance to complement routine phenotypic investigations especially in accurate identification of bacteria and their resistance to antibiotics in resource-limited settings. We noticed a discordance between the phenotypic and genotypic identification of the bacterial isolates. For example, some*. E. coli* and *K. Pneumoniae* were phenotypically misidentified as *Citrobacter spp and an E. cloaca as S. enterica.* Rosenthal et al have posited that bacterial misidentification can arise from laboratory procedural errors, such as culture media contamination or use of suboptimal reagents (e.g., dried agar, expired substrates), which skew biochemical test results (51). Additionally, evolutionary mutations in bacteria, driven by selective pressures like antibiotics or nutrient scarcity, can alter phenotypic changes via genetic mutations or horizontal gene transfer, thereby confounding precise taxonomic identification(52). Misidenfication of bacteria in the clinical setting could result in deleterious clinical outcomes, a phenomenon that can be forestalled by complementary genomic profiling (53,54). Additionally, genomic characterization can also reveal the genetic basis of AMRs even for drugs for which AST were not performed. Also, VFs and plasmid identities from genomic profiling could be used to predict the pathogenicity potential and transmissibility of AMRs respectively(53,55).

ESBL-producing *Enterobacteriaceae* have been implicated in the outbreak of several bacteria borne diseases including food-borne diseases, nosocomial and care-home outbreaks (56–58). In our study, 93% of the previously suspected ESBL producers were confirmed as ESBL producing. Some hospital-based studies in Cape Coast, Ghana (59), and other African countries (10,60,61) have reported similar rates of ESBL producing *Enterobacteriaceae*, especially in *E. coli* and *K. pneumoniae*. However, a lower rate of 14.6% was observed in a hospital-based study in Ontario, Canada (62). Of these ESBL-producing isolates sequenced 19 harboured 17 individual ß-lactamase ARGs of Ambler groups A, B C and D. ß-lactamases associated with ESBLs observed were mainly *SHV, TEM and CTX-M* type genes found in the Ambler group A. These variants are mostly penicillinases and cephalosporinases and tend to be resistant to clavulanate, sulbactam and tazobactam (63,64). This could explain why the addition of clavulanate to ceftazidime did not improve the sensitivity profile during the AST. *CTX-M-15* a member of the Ambler class A was the predominant ß-lactamase ARG among the isolates accounting for 68.4% (13/19). Previous studies in Ghana have reported *CTX-M-15* prevalence of over 80% in *Enterobacteriaceae* (65,66), whereas Asare Yeboah et al (10) reported a lower proportion of 52%. Globally, CTX*-M-15* is the most prevalent ß-lactamase ARG in *Enterobacteriaceae*, with a pooled prevalence reported at 16% which is lower than observed 74% (14/19) in our study. we also observed the presence of oxacillinases including OXA-1, OXA-58 and OXA-402 which also confer ESBL activity (63). Generally, most of the isolates were sensitive to the carbapenems with just above 11% of the isolates (9/79) being phenotypically resistant to meropenem. However, only 2 of these resistant isolates, both *K. pneumoniae* (BC71 and BC75), were part of the whole genome sequenced isolates. These *K. pneumoniae* isolates harboured NDM-1 carbapenemase. NDM-1, an Ambler class B ß-lactamase has previously been described in *E. coli* clinical isolates and also from a hospital environment surveillance study (12,67).

VFs are produced by pathogenic *Enterobacteriaceae* which enable them to colonize, persist, and ultimately cause infections in the host (14,19). Majority of VFs identified in the WGS isolates were in the adherence, effector delivery systems and nutritional or metabolic factors families, mostly in the *E. coli* and *K. pneumoniae.* However, the only *S. enterica* harboured most of the effector delivery systems. Studies conducted in both Ghana and Slovakia (14,15) observed that the prevalence of these 3 VFs among *Enterobacteriaceae*, especially *E*. *coli*, *K. pneumoniae*, *E. cloacae*, and *S*. *enterica* were high. Other studies in Africa and some European countries also corroborate our findings (18,68–70). From our study, the whole genome sequenced isolates included 11 isolates, that is 3 *K. pneumoniae* and 8 *E. coli* which were isolated from urine infections. Generally, for *Enterobacteriaceae* to be able to colonize and cause urinary tract infections depends on their ability to form biofilms and produce several VFs, especially adherence factors (P fimbriae, S fimbriae, type I fimbriae), iron acquisition and sequestration (aerobactin and enterobactins), hemolysin and cytotoxic necrotizing factors (cnf1) (71,72).

Fimbriae are absolute prerequisite for urinary tract colonization and disease causation, because *Enterobacteriaceae* uses that to adhere to host cells and form protective biofilms in the urinary tract contributing to the pathogenesis and persistence of urinary tract infections (73,74). Incidentally, the *E. coli* and *K. pneumoniae* sequenced possessed numerous fimbrial genes including type 1, type 2 (P fimbriae), type 3 and S fimbriae. Specifically, *E. coli* harboured type I fimbriae (*fimA, B, I, D, E G, H, K, F, T, U, V*), P fimbriae (*papB, C, F, G, H, D, F, J, K, X)* and S fimbriae (*sfaD, E, F, C, G, H, S, X, Y)*. Studies in uropathogenic *Enterobacteriaceae* conducted in Benin, Romania, Mongolia, and Egypt reported these fimbrial genes as well (71). Uropathogenic *K. pneumoniae* utilizes two main types of fimbriae for adhesion to the uroepithelium, type 1 and type III (75). However, we observed type II fimbriae (*papF, papK*) in the 3 uropathogenic *K. pneumoniae* isolates in addition to type I fimbriae (*fimA, B, D, E, F, G, H, I, K*), and type III fimbriae (*mrkC, I, F, A, D, B, H, D*).

In addition to these adherence factors the *E. coli* isolates also harboured iron acquisition and sequestration genes including aerobactins (*iucA, iucB, iutA*), and enterobactins (*entA, entC, entE, entF, entS, entB*). The 3 *K. pneumoniae* also possessed an aerobactin (*iutA*.) and enterobactins (*entF, entA, entE, entC, entS*). Siderophores, such as enterobactins and aerobactins, enable uropathogenic *Enterobacteriaceae* to thrive in the host by scavenging iron from host epithelial cells (76). According to Moxley and Payne et al (20,77), *Enterobacteriaceae* requires iron for growth and virulence. Hemolysin (*hlyC, hlyD*), and *cnf1* which the *E. coli* uses to damage cell nutrient and trigger siderophores to sequester irons from host for growth (78) were also observed among the *E. coli* isolates. Nhu et al. reports that hemolysin (*hlyC, D*) and cnf1 were found abundantly in UPEC strains, emphasizing their significance capacity to cause UTI (79)

According to (80,81) most ARGs and VFs are borne on mobile genetic elements, especially plasmids and have become important for the dissemination of these factors within the *Enterobacteriaceae* family through horizontal or vertical gene transfer. Plasmids have been reported as main vehicles for the transmission of ARGs including ESBLs and VF genes in humans and non-human settings (29–31,81). It has been reported that most plasmid replicons found for *Enterobacteriaceae* are I-complex (Incompatibility complex) plasmids and are mostly resistance and fertility plasmids in addition to colicinogenic plasmid (26,29,32,82). In our study, the I-complex we observed most was the IncF (13/19 total Inc plasmids). *IncF* plasmids which are conjugative have been reported in *Enterobacteriaceae* isolated from humans, animal and the environment worldwide (29,83,84). These plasmids have been reported worldwide in the dissemination of AMR genes, especially the ESBLs and particularly *CTX-M-15* (29,83,85). Unsurprisingly, *CTX-M-15* was the commonest ARG observed in our study. In our study, these complex group of plasmid replicons were observed in E*. coli and K. pneumoniae* similar to previous reports (29,83). Other Inc plasmid replicons observed in our study included *IncC, IncH, IncI, IncR* and *IncX*.

The 8 colicins uncovered in our study including *Col (BS512) _1, Col156_1, Col (KPHS6) _1, ColRNAI_1*, *Col440I_1, Col440II_1*, *ColpVC_1* and *Col3M_1* could have been involved in the isolate’s resistance to the antibiotics. Consistent with previous investigations, our study adds to the expanding body of evidence indicating that colicins facilitates AMR dissemination (86–88).

This study revealed the resistant profile and VFs of *Enterobacteriaceae* at CCTH, creating awareness of the serious threat ESBL producing *Enterobacteriaceae* portends to healthcare delivery. It has also highlighted plasmids associated with AMR genes transmission within the *Enterobacteriaceae* populace and informs the importance to using genomic data of bacteria isolates to infer diagnosis and prescriptions.

## Conclusions

Our study revealed high rate of multi-drug AMR among the isolates, particularly to ESBL and narrow spectrum b-lactams. Several ARGs which confer resistance to ESBLs were abundant in the 19 whole sequenced isolates. VFs genes serving as prerequisite for successful Enterobacteriaceae urinary tract colonization and infection were detected in the sequenced isolates. The outcome emphasizes the need to improve routine genomic surveillance to monitor antibiotic resistance trends and guide evidence-based interventions aimed at mitigating the spread of ARE in the Cape Coast Metropolis and other parts of Ghana.

## Limitations

The phenotypic resistance assessment was limited to disc diffusion methods, mainly constrained by logistical limitations. Our findings potentially do not fully capture genetic assessment on the plasmids of the individual isolates due to the usage of illumina short reads which could lead to incomplete reconstructions of plasmids or misassembles. It should be noted that our study circumscribe only those *Enterobacteriaceae* isolates accessible from the bacteriology laboratory at CCTH, and with availability of Socio-demographic data. Thus, our phenotypic AMR assessment do not fully capture available *Enterobacteriaceae* isolates present at the time of this study at the hospital. Also due to funding constrains only 20 isolates out of the 100 isolates were whole genome sequenced; our findings potentially do not fully capture the AMR genotypic assessment of all the retrieved isolates.

## Data Availability

All data produced in the present study are available upon reasonable request to the authors

## Acknowledgements

We extend our heartfelt thanks to Dr. Oheneba Charles Kofi Hagan for the provision of financial support, without which this research would not have been possible. Additionally, we recognize the invaluable contributions of the entire research team, whose dedication and expertise were essential at every stage—from the initial conceptualization and laboratory investigations to the final data analysis and manuscript preparation.

## Authors Contribution

The conceptualization of the study, including the development of the research idea and design of the overall framework, was carried out by B.D., F.H., and O.C.K.H. Microbiology work, such as sample processing, culture, and phenotypic characterization, was performed by B.D., P.M., A.B.M., and A.A.B. Molecular experiments and sequencing procedures, including DNA extraction, PCR, and library preparation, were conducted by B.D. and O.C.K.H. Bioinformatics workflow development and data analysis were undertaken by B.D., and R.O.M. and O.C.K.H., who processed and interpreted the sequencing data. The manuscript was written and revised collaboratively by B.D., F.H.., R.O.M., and O.C.K.H., who contributed to the drafting, critical revision, and final approval of the text. The overall supervision of the project, including oversight of research activities and provision of guidance and resources, was provided by O.C.K.H. and F.H

## Conflict of interests

The authors declare that there are no personal relationships that could have appeared to influence the work reported in this paper

## Notes

### Competing Interest Statement

The authors have declared no competing interest.

### Funding Statement

This study did not receive any funding

### Author Declarations

Cape Coast teaching Hospital Ethical Clearance Review Committee (CCTHERC) of Cape Coast Teaching Hospital gave the ethical approval with number CCTHERC/EC/2023/102.

### Summary of Updates

This version of the manuscript has been revised to update the some references that still appeared as APA to Vancouver, and to also show the full view of fig1.

