## Additional File 1 for "Phenotypic and Genotypic Profile of Enterobacteriaceae Isolated at a Teaching Hospital in Ghana"

**Additional File 1**: Phenotypic and genotypic (MLST, rMLST) classification of Isolates

| ID. | Phenotypic ID | rMLST | MLST | Specimen Source | Department |
| --- | --- | --- | --- | --- | --- |
| BC04 | *E. coli* | *E. coli* | *E. coli* | Urine | FSW |
| BC06 | *S. enterica* | *E. cloacae* | *E. cloacae* | Stool | OPD |
| BC15 | *E. coli* | *E. coli* | *E. coli* | HVS | OPD |
| BC16 | *K. Pneumoniae* | *K. Pneumoniae* | *K. Pneumoniae* | Sputum | FMW |
| BC18 | *E. coli* | *E. coli* | *E. coli* | Urine | FMW |
| BC20 | *Citrobacter spp* | *K. Pneumoniae* | *K. Pneumoniae* | Urine | O/G |
| BC26 | *E. coli* | *E. coli* | *E. coli* | Urine | OPD |
| BC35 | *E. cloacae* | *E. cloacae* | *E. cloacae* | Urine | MMW |
| BC37 | *K. Pneumoniae* | *K. Pneumoniae* | *K. Pneumoniae* | Urine | O/G |
| BC38 | *S. enterica* | *S. enterica* | *S. enterica* | Blood | PMW |
| BC54 | *K. Pneumoniae* | *K. Pneumoniae* | *K. Pneumoniae* | Wound | MMW |
| BC64 | *E. coli* | *E. coli* | *E. coli* | HVS | OPD |
| BC71 | *Citrobacter spp* | *E. cloacae* | *K. Pneumoniae* | Urine | A/E |
| BC73 | *E. coli* | *E. coli* | *E. coli* | Urine | OPD |
| BC75 | *K. Pneumoniae* | *K. Pneumoniae* | *K. Pneumoniae* | HVS | OPD |
| BC76 | Proteous mirabilis | *K. Pneumoniae* | *K. Pneumoniae* | Wound | A/E |
| BC77 | *E. coli* | *P. aeruginosa* | *P. aeruginosa* | Urine | MSW |
| BC81 | *Citrobacter spp* | *Acinetobacter baumanni* | *E. coli* | Urine | OPD |
| BC82 | *Citrobacter spp* | *Acinetobacter baumanni* | *E. coli* | Urine | OPD |
| BC94 | *E. coli* | *K. Pneumoniae* | *K. Pneumoniae* | Pleural Aspirate | ICU |
