## Additional File 2 for "Phenotypic and Genotypic Profile of Enterobacteriaceae Isolated at a Teaching Hospital in Ghana"

**Additional File 2:** Ambler Classification of β-lactamase Genes Identified in Isolates

| **Class** | **ARGs** | ***Escherichia coli*** | | | | | | | | ***Klebsiella Pneumoniae*** | | | | | | | | ***Enterobacter cloacae*** | | ***Salmonella enterica*** | **Number of Isolates** |
| --- | --- | --- | --- | --- | --- | --- | --- | --- | --- | --- | --- | --- | --- | --- | --- | --- | --- | --- | --- | --- | --- |
|  |  | **BC04** | **BC15** | **BC18** | **BC26** | **BC64** | **BC73** | **BC81** | **BC82** | **BC16** | **BC20** | **BC37** | **BC54** | **BC71** | **BC75** | **BC76** | **BC94** | **BC06** | **BC35** | **BC38** |  |
| A | SHV-106 |  |  |  |  |  |  |  |  |  |  |  |  |  |  |  |  |  |  |  | **4** |
|  | SHV-80 |  |  |  |  |  |  |  |  |  |  |  |  |  |  |  |  |  |  |  | **1** |
|  | SHV-187 |  |  |  |  |  |  |  |  |  |  |  |  |  |  |  |  |  |  |  | **3** |
|  | TEM-1 |  |  |  |  |  |  |  |  |  |  |  |  |  |  |  |  |  |  |  | **6** |
|  | TEM-35 |  |  |  |  |  |  |  |  |  |  |  |  |  |  |  |  |  |  |  | **1** |
|  | CTX-M-15 |  |  |  |  |  |  |  |  |  |  |  |  |  |  |  |  |  |  |  | **13** |
| B | NDM-1 |  |  |  |  |  |  |  |  |  |  |  |  |  |  |  |  |  |  |  | **4** |
| C | ADC-169 |  |  |  |  |  |  |  |  |  |  |  |  |  |  |  |  |  |  |  | **2** |
|  | Escherichia_coli_ampC_beta-lactamase |  |  |  |  |  |  |  |  |  |  |  |  |  |  |  |  |  |  |  | **3** |
|  | EC-5 |  |  |  |  |  |  |  |  |  |  |  |  |  |  |  |  |  |  |  | **4** |
|  | EC-15 |  |  |  |  |  |  |  |  |  |  |  |  |  |  |  |  |  |  |  | **1** |
|  | CMH-3 |  |  |  |  |  |  |  |  |  |  |  |  |  |  |  |  |  |  |  | **1** |
|  | CMH-6 |  |  |  |  |  |  |  |  |  |  |  |  |  |  |  |  |  |  |  | **2** |
|  | CMY-59 |  |  |  |  |  |  |  |  |  |  |  |  |  |  |  |  |  |  |  | **1** |
| D | OXA-1 |  |  |  |  |  |  |  |  |  |  |  |  |  |  |  |  |  |  |  | **8** |
|  | OXA-402 |  |  |  |  |  |  |  |  |  |  |  |  |  |  |  |  |  |  |  | **2** |
|  | OXA-58 |  |  |  |  |  |  |  |  |  |  |  |  |  |  |  |  |  |  |  | **2** |
| Total | | **2** | **2** | **2** | **3** | **2** | **3** | **8** | **7** | **5** | **2** | **6** | **3** | **7** | **6** | **1** | **1** | **1** | **2** | **0** | ***** |

This table presents the β-lactamase antibiotic resistance genes (ARGs) identified in Escherichia coli, Klebsiella pneumoniae, Enterobacter cloacae, and Salmonella enterica isolates, categorized according to Ambler classes A–D. Each column (BC04–BC38) represents a distinct bacterial isolate, while rows correspond to individual ARG variants. The presence of a gene in an isolate is indicated by a dark-blue–shaded cell. Red-shaded cells highlight ARGs that are rarely reported among Enterobacteriaceae in Ghana. The rightmost column indicates the total number of isolates harboring each gene, and the bottom row shows the total number of ARGs detected per isolate
