## Additional File 3 for "Phenotypic and Genotypic Profile of Enterobacteriaceae Isolated at a Teaching Hospital in Ghana"

**Additional File 3**: Table of Virulence Factors profiled in the *Enterobacteriaceae* Isolates

| **VFs** | ***Escherichia coli*** | | | | | | | | ***Klebsiella Pneumoniae*** | | | | | | | | ***Enterobacter cloacae*** | | ***Salmonella enterica*** | **Number of Isolates** |
| --- | --- | --- | --- | --- | --- | --- | --- | --- | --- | --- | --- | --- | --- | --- | --- | --- | --- | --- | --- | --- |
|  | **BC04** | **BC15** | **BC18** | **BC26** | **BC64** | **BC73** | **BC81** | **BC82** | **BC16** | **BC20** | **BC37** | **BC54** | **BC71** | **BC75** | **BC76** | **BC94** | **BC06** | **BC35** | **BC38** |  |
| abaI |  |  |  |  |  |  |  |  |  |  |  |  |  |  |  |  |  |  |  | **2** |
| abaR |  |  |  |  |  |  |  |  |  |  |  |  |  |  |  |  |  |  |  | **2** |
| ACICU |  |  |  |  |  |  |  |  |  |  |  |  |  |  |  |  |  |  |  | **2** |
| acrA |  |  |  |  |  |  |  |  |  |  |  |  |  |  |  |  |  |  |  | **7** |
| acrB |  |  |  |  |  |  |  |  |  |  |  |  |  |  |  |  |  |  |  | **7** |
| adeF |  |  |  |  |  |  |  |  |  |  |  |  |  |  |  |  |  |  |  | **2** |
| adeG |  |  |  |  |  |  |  |  |  |  |  |  |  |  |  |  |  |  |  | **2** |
| adeH |  |  |  |  |  |  |  |  |  |  |  |  |  |  |  |  |  |  |  | **2** |
| aslA |  |  |  |  |  |  |  |  |  |  |  |  |  |  |  |  |  |  |  | **4** |
| ata |  |  |  |  |  |  |  |  |  |  |  |  |  |  |  |  |  |  |  | **2** |
| avrA |  |  |  |  |  |  |  |  |  |  |  |  |  |  |  |  |  |  |  | **1** |
| barA |  |  |  |  |  |  |  |  |  |  |  |  |  |  |  |  |  |  |  | **2** |
| barB |  |  |  |  |  |  |  |  |  |  |  |  |  |  |  |  |  |  |  | **2** |
| basA |  |  |  |  |  |  |  |  |  |  |  |  |  |  |  |  |  |  |  | **2** |
| basB |  |  |  |  |  |  |  |  |  |  |  |  |  |  |  |  |  |  |  | **2** |
| basC |  |  |  |  |  |  |  |  |  |  |  |  |  |  |  |  |  |  |  | **2** |
| basD |  |  |  |  |  |  |  |  |  |  |  |  |  |  |  |  |  |  |  | **2** |
| basF |  |  |  |  |  |  |  |  |  |  |  |  |  |  |  |  |  |  |  | **2** |
| basG |  |  |  |  |  |  |  |  |  |  |  |  |  |  |  |  |  |  |  | **2** |
| bash |  |  |  |  |  |  |  |  |  |  |  |  |  |  |  |  |  |  |  | **2** |
| basI |  |  |  |  |  |  |  |  |  |  |  |  |  |  |  |  |  |  |  | **2** |
| basJ |  |  |  |  |  |  |  |  |  |  |  |  |  |  |  |  |  |  |  | **2** |
| bauB |  |  |  |  |  |  |  |  |  |  |  |  |  |  |  |  |  |  |  | **2** |
| bauC |  |  |  |  |  |  |  |  |  |  |  |  |  |  |  |  |  |  |  | **2** |
| bauD |  |  |  |  |  |  |  |  |  |  |  |  |  |  |  |  |  |  |  | **2** |
| bauE |  |  |  |  |  |  |  |  |  |  |  |  |  |  |  |  |  |  |  | **2** |
| bauF |  |  |  |  |  |  |  |  |  |  |  |  |  |  |  |  |  |  |  | **2** |
| cap8L |  |  |  |  |  |  |  |  |  |  |  |  |  |  |  |  |  |  |  | **2** |
| cap8O |  |  |  |  |  |  |  |  |  |  |  |  |  |  |  |  |  |  |  | **1** |
| cfaA |  |  |  |  |  |  |  |  |  |  |  |  |  |  |  |  |  |  |  | **1** |
| cfaB |  |  |  |  |  |  |  |  |  |  |  |  |  |  |  |  |  |  |  | **1** |
| cfaC |  |  |  |  |  |  |  |  |  |  |  |  |  |  |  |  |  |  |  | **1** |
| cfaD/cfaE |  |  |  |  |  |  |  |  |  |  |  |  |  |  |  |  |  |  |  | **1** |
| cgsD |  |  |  |  |  |  |  |  |  |  |  |  |  |  |  |  |  |  |  | **10** |
| cgsE |  |  |  |  |  |  |  |  |  |  |  |  |  |  |  |  |  |  |  | **9** |
| cgsF |  |  |  |  |  |  |  |  |  |  |  |  |  |  |  |  |  |  |  | **10** |
| cgsG |  |  |  |  |  |  |  |  |  |  |  |  |  |  |  |  |  |  |  | **10** |
| chuA |  |  |  |  |  |  |  |  |  |  |  |  |  |  |  |  |  |  |  | **4** |
| chuS |  |  |  |  |  |  |  |  |  |  |  |  |  |  |  |  |  |  |  | **4** |
| chuT |  |  |  |  |  |  |  |  |  |  |  |  |  |  |  |  |  |  |  | **4** |
| chuU |  |  |  |  |  |  |  |  |  |  |  |  |  |  |  |  |  |  |  | **4** |
| chuV |  |  |  |  |  |  |  |  |  |  |  |  |  |  |  |  |  |  |  | **4** |
| chuW |  |  |  |  |  |  |  |  |  |  |  |  |  |  |  |  |  |  |  | **4** |
| chuX |  |  |  |  |  |  |  |  |  |  |  |  |  |  |  |  |  |  |  | **4** |
| chuY |  |  |  |  |  |  |  |  |  |  |  |  |  |  |  |  |  |  |  | **4** |
| clbA |  |  |  |  |  |  |  |  |  |  |  |  |  |  |  |  |  |  |  | **2** |
| clbB |  |  |  |  |  |  |  |  |  |  |  |  |  |  |  |  |  |  |  | **2** |
| clbC |  |  |  |  |  |  |  |  |  |  |  |  |  |  |  |  |  |  |  | **2** |
| clbD |  |  |  |  |  |  |  |  |  |  |  |  |  |  |  |  |  |  |  | **2** |
| clbE |  |  |  |  |  |  |  |  |  |  |  |  |  |  |  |  |  |  |  | **2** |
| clbF |  |  |  |  |  |  |  |  |  |  |  |  |  |  |  |  |  |  |  | **2** |
| clbG |  |  |  |  |  |  |  |  |  |  |  |  |  |  |  |  |  |  |  | **2** |
| clbH |  |  |  |  |  |  |  |  |  |  |  |  |  |  |  |  |  |  |  | **2** |
| clbI |  |  |  |  |  |  |  |  |  |  |  |  |  |  |  |  |  |  |  | **2** |
| clbJ |  |  |  |  |  |  |  |  |  |  |  |  |  |  |  |  |  |  |  | **2** |
| clbL |  |  |  |  |  |  |  |  |  |  |  |  |  |  |  |  |  |  |  | **2** |
| clbM |  |  |  |  |  |  |  |  |  |  |  |  |  |  |  |  |  |  |  | **2** |
| clbN |  |  |  |  |  |  |  |  |  |  |  |  |  |  |  |  |  |  |  | **2** |
| clbO |  |  |  |  |  |  |  |  |  |  |  |  |  |  |  |  |  |  |  | **2** |
| clbP |  |  |  |  |  |  |  |  |  |  |  |  |  |  |  |  |  |  |  | **2** |
| clbQ |  |  |  |  |  |  |  |  |  |  |  |  |  |  |  |  |  |  |  | **2** |
| clbS |  |  |  |  |  |  |  |  |  |  |  |  |  |  |  |  |  |  |  | **2** |
| clpV/tssH |  |  |  |  |  |  |  |  |  |  |  |  |  |  |  |  |  |  |  | **10** |
| cnf1 |  |  |  |  |  |  |  |  |  |  |  |  |  |  |  |  |  |  |  | **2** |
| csgA |  |  |  |  |  |  |  |  |  |  |  |  |  |  |  |  |  |  |  | **4** |
| csgB |  |  |  |  |  |  |  |  |  |  |  |  |  |  |  |  |  |  |  | **9** |
| csgC |  |  |  |  |  |  |  |  |  |  |  |  |  |  |  |  |  |  |  | **9** |
| csuA |  |  |  |  |  |  |  |  |  |  |  |  |  |  |  |  |  |  |  | **2** |
| csuA/B |  |  |  |  |  |  |  |  |  |  |  |  |  |  |  |  |  |  |  | **2** |
| csuB |  |  |  |  |  |  |  |  |  |  |  |  |  |  |  |  |  |  |  | **2** |
| csuC |  |  |  |  |  |  |  |  |  |  |  |  |  |  |  |  |  |  |  | **2** |
| csuD |  |  |  |  |  |  |  |  |  |  |  |  |  |  |  |  |  |  |  | **2** |
| csuE |  |  |  |  |  |  |  |  |  |  |  |  |  |  |  |  |  |  |  | **2** |
| dotU/tssL |  |  |  |  |  |  |  |  |  |  |  |  |  |  |  |  |  |  |  | **8** |
| draA |  |  |  |  |  |  |  |  |  |  |  |  |  |  |  |  |  |  |  | **1** |
| draB |  |  |  |  |  |  |  |  |  |  |  |  |  |  |  |  |  |  |  | **1** |
| draC |  |  |  |  |  |  |  |  |  |  |  |  |  |  |  |  |  |  |  | **1** |
| draD |  |  |  |  |  |  |  |  |  |  |  |  |  |  |  |  |  |  |  | **1** |
| draP |  |  |  |  |  |  |  |  |  |  |  |  |  |  |  |  |  |  |  | **1** |
| entA |  |  |  |  |  |  |  |  |  |  |  |  |  |  |  |  |  |  |  | **16** |
| entB |  |  |  |  |  |  |  |  |  |  |  |  |  |  |  |  |  |  |  | **16** |
| entC |  |  |  |  |  |  |  |  |  |  |  |  |  |  |  |  |  |  |  | **15** |
| entD |  |  |  |  |  |  |  |  |  |  |  |  |  |  |  |  |  |  |  | **15** |
| entE |  |  |  |  |  |  |  |  |  |  |  |  |  |  |  |  |  |  |  | **15** |
| entF |  |  |  |  |  |  |  |  |  |  |  |  |  |  |  |  |  |  |  | **15** |
| entS |  |  |  |  |  |  |  |  |  |  |  |  |  |  |  |  |  |  |  | **15** |
| espL1 |  |  |  |  |  |  |  |  |  |  |  |  |  |  |  |  |  |  |  | **3** |
| espL4 |  |  |  |  |  |  |  |  |  |  |  |  |  |  |  |  |  |  |  | **4** |
| espX1 |  |  |  |  |  |  |  |  |  |  |  |  |  |  |  |  |  |  |  | **2** |
| espX5 |  |  |  |  |  |  |  |  |  |  |  |  |  |  |  |  |  |  |  | **5** |
| esxC |  |  |  |  |  |  |  |  |  |  |  |  |  |  |  |  |  |  |  | **1** |
| esxD |  |  |  |  |  |  |  |  |  |  |  |  |  |  |  |  |  |  |  | **1** |
| fdeC |  |  |  |  |  |  |  |  |  |  |  |  |  |  |  |  |  |  |  | **4** |
| fepA |  |  |  |  |  |  |  |  |  |  |  |  |  |  |  |  |  |  |  | **15** |
| fepB |  |  |  |  |  |  |  |  |  |  |  |  |  |  |  |  |  |  |  | **15** |
| fepC |  |  |  |  |  |  |  |  |  |  |  |  |  |  |  |  |  |  |  | **16** |
| fepD |  |  |  |  |  |  |  |  |  |  |  |  |  |  |  |  |  |  |  | **15** |
| fepE |  |  |  |  |  |  |  |  |  |  |  |  |  |  |  |  |  |  |  | **12** |
| fepG |  |  |  |  |  |  |  |  |  |  |  |  |  |  |  |  |  |  |  | **12** |
| fes |  |  |  |  |  |  |  |  |  |  |  |  |  |  |  |  |  |  |  | **16** |
| fha |  |  |  |  |  |  |  |  |  |  |  |  |  |  |  |  |  |  |  | **3** |
| fimA |  |  |  |  |  |  |  |  |  |  |  |  |  |  |  |  |  |  |  | **9** |
| fimB |  |  |  |  |  |  |  |  |  |  |  |  |  |  |  |  |  |  |  | **16** |
| fimC |  |  |  |  |  |  |  |  |  |  |  |  |  |  |  |  |  |  |  | **16** |
| fimD |  |  |  |  |  |  |  |  |  |  |  |  |  |  |  |  |  |  |  | **16** |
| fimE |  |  |  |  |  |  |  |  |  |  |  |  |  |  |  |  |  |  |  | **17** |
| fimF |  |  |  |  |  |  |  |  |  |  |  |  |  |  |  |  |  |  |  | **17** |
| fimG |  |  |  |  |  |  |  |  |  |  |  |  |  |  |  |  |  |  |  | **16** |
| fimH |  |  |  |  |  |  |  |  |  |  |  |  |  |  |  |  |  |  |  | **16** |
| fimI |  |  |  |  |  |  |  |  |  |  |  |  |  |  |  |  |  |  |  | **16** |
| fimK |  |  |  |  |  |  |  |  |  |  |  |  |  |  |  |  |  |  |  | **7** |
| fimT |  |  |  |  |  |  |  |  |  |  |  |  |  |  |  |  |  |  |  | **2** |
| fimU |  |  |  |  |  |  |  |  |  |  |  |  |  |  |  |  |  |  |  | **2** |
| fimV |  |  |  |  |  |  |  |  |  |  |  |  |  |  |  |  |  |  |  | **2** |
| fimW |  |  |  |  |  |  |  |  |  |  |  |  |  |  |  |  |  |  |  | **1** |
| fimY |  |  |  |  |  |  |  |  |  |  |  |  |  |  |  |  |  |  |  | **1** |
| fimZ |  |  |  |  |  |  |  |  |  |  |  |  |  |  |  |  |  |  |  | **1** |
| focA |  |  |  |  |  |  |  |  |  |  |  |  |  |  |  |  |  |  |  | **1** |
| focC |  |  |  |  |  |  |  |  |  |  |  |  |  |  |  |  |  |  |  | **1** |
| focD |  |  |  |  |  |  |  |  |  |  |  |  |  |  |  |  |  |  |  | **1** |
| focF |  |  |  |  |  |  |  |  |  |  |  |  |  |  |  |  |  |  |  | **1** |
| focG |  |  |  |  |  |  |  |  |  |  |  |  |  |  |  |  |  |  |  | **1** |
| focH |  |  |  |  |  |  |  |  |  |  |  |  |  |  |  |  |  |  |  | **1** |
| focI |  |  |  |  |  |  |  |  |  |  |  |  |  |  |  |  |  |  |  | **1** |
| fur |  |  |  |  |  |  |  |  |  |  |  |  |  |  |  |  |  |  |  | **1** |
| fyuA/psn |  |  |  |  |  |  |  |  |  |  |  |  |  |  |  |  |  |  |  | **12** |
| galF |  |  |  |  |  |  |  |  |  |  |  |  |  |  |  |  |  |  |  | **10** |
| gndA |  |  |  |  |  |  |  |  |  |  |  |  |  |  |  |  |  |  |  | **4** |
| gspC |  |  |  |  |  |  |  |  |  |  |  |  |  |  |  |  |  |  |  | **4** |
| gspD |  |  |  |  |  |  |  |  |  |  |  |  |  |  |  |  |  |  |  | **4** |
| gspE |  |  |  |  |  |  |  |  |  |  |  |  |  |  |  |  |  |  |  | **3** |
| gspE1 |  |  |  |  |  |  |  |  |  |  |  |  |  |  |  |  |  |  |  | **2** |
| gspE2 |  |  |  |  |  |  |  |  |  |  |  |  |  |  |  |  |  |  |  | **2** |
| gspF |  |  |  |  |  |  |  |  |  |  |  |  |  |  |  |  |  |  |  | **3** |
| gspG |  |  |  |  |  |  |  |  |  |  |  |  |  |  |  |  |  |  |  | **4** |
| gspH |  |  |  |  |  |  |  |  |  |  |  |  |  |  |  |  |  |  |  | **6** |
| gspI |  |  |  |  |  |  |  |  |  |  |  |  |  |  |  |  |  |  |  | **3** |
| gspJ |  |  |  |  |  |  |  |  |  |  |  |  |  |  |  |  |  |  |  | **3** |
| gspK |  |  |  |  |  |  |  |  |  |  |  |  |  |  |  |  |  |  |  | **3** |
| gspL |  |  |  |  |  |  |  |  |  |  |  |  |  |  |  |  |  |  |  | **7** |
| gspM |  |  |  |  |  |  |  |  |  |  |  |  |  |  |  |  |  |  |  | **4** |
| gspN |  |  |  |  |  |  |  |  |  |  |  |  |  |  |  |  |  |  |  | **2** |
| gspO/pilD |  |  |  |  |  |  |  |  |  |  |  |  |  |  |  |  |  |  |  | **2** |
| hcp/tssD |  |  |  |  |  |  |  |  |  |  |  |  |  |  |  |  |  |  |  | **7** |
| hcp1/tssD1 |  |  |  |  |  |  |  |  |  |  |  |  |  |  |  |  |  |  |  | **5** |
| hcp2/tssD2 |  |  |  |  |  |  |  |  |  |  |  |  |  |  |  |  |  |  |  | **1** |
| hilA |  |  |  |  |  |  |  |  |  |  |  |  |  |  |  |  |  |  |  | **1** |
| hilC |  |  |  |  |  |  |  |  |  |  |  |  |  |  |  |  |  |  |  | **1** |
| hilD |  |  |  |  |  |  |  |  |  |  |  |  |  |  |  |  |  |  |  | **1** |
| hld |  |  |  |  |  |  |  |  |  |  |  |  |  |  |  |  |  |  |  | **1** |
| hlyA |  |  |  |  |  |  |  |  |  |  |  |  |  |  |  |  |  |  |  | **2** |
| hlyB |  |  |  |  |  |  |  |  |  |  |  |  |  |  |  |  |  |  |  | **2** |
| hlyC |  |  |  |  |  |  |  |  |  |  |  |  |  |  |  |  |  |  |  | **2** |
| hlyD |  |  |  |  |  |  |  |  |  |  |  |  |  |  |  |  |  |  |  | **2** |
| iacP |  |  |  |  |  |  |  |  |  |  |  |  |  |  |  |  |  |  |  | **1** |
| iagB |  |  |  |  |  |  |  |  |  |  |  |  |  |  |  |  |  |  |  | **1** |
| ibeB |  |  |  |  |  |  |  |  |  |  |  |  |  |  |  |  |  |  |  | **9** |
| ibeC |  |  |  |  |  |  |  |  |  |  |  |  |  |  |  |  |  |  |  | **9** |
| icmF/tssM |  |  |  |  |  |  |  |  |  |  |  |  |  |  |  |  |  |  |  | **3** |
| impA/tssA |  |  |  |  |  |  |  |  |  |  |  |  |  |  |  |  |  |  |  | **4** |
| invA |  |  |  |  |  |  |  |  |  |  |  |  |  |  |  |  |  |  |  | **1** |
| invB |  |  |  |  |  |  |  |  |  |  |  |  |  |  |  |  |  |  |  | **1** |
| invC/sctN |  |  |  |  |  |  |  |  |  |  |  |  |  |  |  |  |  |  |  | **1** |
| invE |  |  |  |  |  |  |  |  |  |  |  |  |  |  |  |  |  |  |  | **1** |
| invF |  |  |  |  |  |  |  |  |  |  |  |  |  |  |  |  |  |  |  | **1** |
| invG |  |  |  |  |  |  |  |  |  |  |  |  |  |  |  |  |  |  |  | **1** |
| invH |  |  |  |  |  |  |  |  |  |  |  |  |  |  |  |  |  |  |  | **1** |
| invI |  |  |  |  |  |  |  |  |  |  |  |  |  |  |  |  |  |  |  | **1** |
| invJ |  |  |  |  |  |  |  |  |  |  |  |  |  |  |  |  |  |  |  | **1** |
| ipfA |  |  |  |  |  |  |  |  |  |  |  |  |  |  |  |  |  |  |  | **1** |
| ipfB |  |  |  |  |  |  |  |  |  |  |  |  |  |  |  |  |  |  |  | **1** |
| ipfC |  |  |  |  |  |  |  |  |  |  |  |  |  |  |  |  |  |  |  | **1** |
| ipfE |  |  |  |  |  |  |  |  |  |  |  |  |  |  |  |  |  |  |  | **1** |
| iroB |  |  |  |  |  |  |  |  |  |  |  |  |  |  |  |  |  |  |  | **2** |
| iroC |  |  |  |  |  |  |  |  |  |  |  |  |  |  |  |  |  |  |  | **2** |
| iroD |  |  |  |  |  |  |  |  |  |  |  |  |  |  |  |  |  |  |  | **2** |
| iroE |  |  |  |  |  |  |  |  |  |  |  |  |  |  |  |  |  |  |  | **8** |
| iroN |  |  |  |  |  |  |  |  |  |  |  |  |  |  |  |  |  |  |  | **2** |
| irp1 |  |  |  |  |  |  |  |  |  |  |  |  |  |  |  |  |  |  |  | **12** |
| irp2 |  |  |  |  |  |  |  |  |  |  |  |  |  |  |  |  |  |  |  | **12** |
| isdl |  |  |  |  |  |  |  |  |  |  |  |  |  |  |  |  |  |  |  | **1** |
| iucA |  |  |  |  |  |  |  |  |  |  |  |  |  |  |  |  |  |  |  | **6** |
| iucB |  |  |  |  |  |  |  |  |  |  |  |  |  |  |  |  |  |  |  | **7** |
| iucC |  |  |  |  |  |  |  |  |  |  |  |  |  |  |  |  |  |  |  | **7** |
| iucD |  |  |  |  |  |  |  |  |  |  |  |  |  |  |  |  |  |  |  | **7** |
| iutA |  |  |  |  |  |  |  |  |  |  |  |  |  |  |  |  |  |  |  | **13** |
| KP1_RS17220 |  |  |  |  |  |  |  |  |  |  |  |  |  |  |  |  |  |  |  | **3** |
| KP1_RS17225 |  |  |  |  |  |  |  |  |  |  |  |  |  |  |  |  |  |  |  | **5** |
| KP1_RS17230 |  |  |  |  |  |  |  |  |  |  |  |  |  |  |  |  |  |  |  | **6** |
| KP1_RS17240 |  |  |  |  |  |  |  |  |  |  |  |  |  |  |  |  |  |  |  | **5** |
| KP1_RS17280 |  |  |  |  |  |  |  |  |  |  |  |  |  |  |  |  |  |  |  | **5** |
| KP1_RS17355 |  |  |  |  |  |  |  |  |  |  |  |  |  |  |  |  |  |  |  | **4** |
| kpsC |  |  |  |  |  |  |  |  |  |  |  |  |  |  |  |  |  |  |  | **4** |
| kpsD |  |  |  |  |  |  |  |  |  |  |  |  |  |  |  |  |  |  |  | **4** |
| kpsE |  |  |  |  |  |  |  |  |  |  |  |  |  |  |  |  |  |  |  | **4** |
| kpsF |  |  |  |  |  |  |  |  |  |  |  |  |  |  |  |  |  |  |  | **4** |
| kpsM |  |  |  |  |  |  |  |  |  |  |  |  |  |  |  |  |  |  |  | **2** |
| kpsS |  |  |  |  |  |  |  |  |  |  |  |  |  |  |  |  |  |  |  | **4** |
| kpsU |  |  |  |  |  |  |  |  |  |  |  |  |  |  |  |  |  |  |  | **3** |
| lpsB |  |  |  |  |  |  |  |  |  |  |  |  |  |  |  |  |  |  |  | **2** |
| lpxA |  |  |  |  |  |  |  |  |  |  |  |  |  |  |  |  |  |  |  | **2** |
| lpxB |  |  |  |  |  |  |  |  |  |  |  |  |  |  |  |  |  |  |  | **2** |
| lpxC |  |  |  |  |  |  |  |  |  |  |  |  |  |  |  |  |  |  |  | **2** |
| lpxD |  |  |  |  |  |  |  |  |  |  |  |  |  |  |  |  |  |  |  | **2** |
| lpxL |  |  |  |  |  |  |  |  |  |  |  |  |  |  |  |  |  |  |  | **2** |
| lpxM |  |  |  |  |  |  |  |  |  |  |  |  |  |  |  |  |  |  |  | **2** |
| mgtB |  |  |  |  |  |  |  |  |  |  |  |  |  |  |  |  |  |  |  | **1** |
| mgtC |  |  |  |  |  |  |  |  |  |  |  |  |  |  |  |  |  |  |  | **1** |
| Mig-14 |  |  |  |  |  |  |  |  |  |  |  |  |  |  |  |  |  |  |  | **1** |
| misL |  |  |  |  |  |  |  |  |  |  |  |  |  |  |  |  |  |  |  | **1** |
| mrkA |  |  |  |  |  |  |  |  |  |  |  |  |  |  |  |  |  |  |  | **6** |
| mrkB |  |  |  |  |  |  |  |  |  |  |  |  |  |  |  |  |  |  |  | **6** |
| mrkC |  |  |  |  |  |  |  |  |  |  |  |  |  |  |  |  |  |  |  | **6** |
| mrkD |  |  |  |  |  |  |  |  |  |  |  |  |  |  |  |  |  |  |  | **6** |
| mrkF |  |  |  |  |  |  |  |  |  |  |  |  |  |  |  |  |  |  |  | **7** |
| mrkH |  |  |  |  |  |  |  |  |  |  |  |  |  |  |  |  |  |  |  | **7** |
| mrkI |  |  |  |  |  |  |  |  |  |  |  |  |  |  |  |  |  |  |  | **7** |
| mrkJ |  |  |  |  |  |  |  |  |  |  |  |  |  |  |  |  |  |  |  | **6** |
| ompA |  |  |  |  |  |  |  |  |  |  |  |  |  |  |  |  |  |  |  | **15** |
| orgA/sctK |  |  |  |  |  |  |  |  |  |  |  |  |  |  |  |  |  |  |  | **1** |
| orgB/SctL |  |  |  |  |  |  |  |  |  |  |  |  |  |  |  |  |  |  |  | **1** |
| orgC |  |  |  |  |  |  |  |  |  |  |  |  |  |  |  |  |  |  |  | **1** |
| papB |  |  |  |  |  |  |  |  |  |  |  |  |  |  |  |  |  |  |  | **4** |
| papC |  |  |  |  |  |  |  |  |  |  |  |  |  |  |  |  |  |  |  | **4** |
| papD |  |  |  |  |  |  |  |  |  |  |  |  |  |  |  |  |  |  |  | **4** |
| papE |  |  |  |  |  |  |  |  |  |  |  |  |  |  |  |  |  |  |  | **1** |
| papF |  |  |  |  |  |  |  |  |  |  |  |  |  |  |  |  |  |  |  | **5** |
| papG |  |  |  |  |  |  |  |  |  |  |  |  |  |  |  |  |  |  |  | **5** |
| papH |  |  |  |  |  |  |  |  |  |  |  |  |  |  |  |  |  |  |  | **2** |
| papI |  |  |  |  |  |  |  |  |  |  |  |  |  |  |  |  |  |  |  | **2** |
| papJ |  |  |  |  |  |  |  |  |  |  |  |  |  |  |  |  |  |  |  | **7** |
| papK |  |  |  |  |  |  |  |  |  |  |  |  |  |  |  |  |  |  |  | **6** |
| papX |  |  |  |  |  |  |  |  |  |  |  |  |  |  |  |  |  |  |  | **8** |
| pbpG |  |  |  |  |  |  |  |  |  |  |  |  |  |  |  |  |  |  |  |  |
| pgaA |  |  |  |  |  |  |  |  |  |  |  |  |  |  |  |  |  |  |  | **2** |
| pgaB |  |  |  |  |  |  |  |  |  |  |  |  |  |  |  |  |  |  |  | **2** |
| pgaC |  |  |  |  |  |  |  |  |  |  |  |  |  |  |  |  |  |  |  | **2** |
| pgaD |  |  |  |  |  |  |  |  |  |  |  |  |  |  |  |  |  |  |  | **2** |
| phoP |  |  |  |  |  |  |  |  |  |  |  |  |  |  |  |  |  |  |  | **1** |
| phoQ |  |  |  |  |  |  |  |  |  |  |  |  |  |  |  |  |  |  |  | **1** |
| pipB |  |  |  |  |  |  |  |  |  |  |  |  |  |  |  |  |  |  |  | **1** |
| pipB2 |  |  |  |  |  |  |  |  |  |  |  |  |  |  |  |  |  |  |  | **1** |
| pmrA |  |  |  |  |  |  |  |  |  |  |  |  |  |  |  |  |  |  |  | **1** |
| pmrB |  |  |  |  |  |  |  |  |  |  |  |  |  |  |  |  |  |  |  | **1** |
| prgH |  |  |  |  |  |  |  |  |  |  |  |  |  |  |  |  |  |  |  | **1** |
| Prgl |  |  |  |  |  |  |  |  |  |  |  |  |  |  |  |  |  |  |  | **1** |
| prgJ |  |  |  |  |  |  |  |  |  |  |  |  |  |  |  |  |  |  |  | **1** |
| prgK |  |  |  |  |  |  |  |  |  |  |  |  |  |  |  |  |  |  |  | **1** |
| pilB |  |  |  |  |  |  |  |  |  |  |  |  |  |  |  |  |  |  |  | **2** |
| pilC |  |  |  |  |  |  |  |  |  |  |  |  |  |  |  |  |  |  |  | **2** |
| pilF |  |  |  |  |  |  |  |  |  |  |  |  |  |  |  |  |  |  |  | **2** |
| pilG |  |  |  |  |  |  |  |  |  |  |  |  |  |  |  |  |  |  |  | **2** |
| pilH |  |  |  |  |  |  |  |  |  |  |  |  |  |  |  |  |  |  |  | **2** |
| pilI |  |  |  |  |  |  |  |  |  |  |  |  |  |  |  |  |  |  |  | **2** |
| pilJ |  |  |  |  |  |  |  |  |  |  |  |  |  |  |  |  |  |  |  | **2** |
| pilM |  |  |  |  |  |  |  |  |  |  |  |  |  |  |  |  |  |  |  | **2** |
| pilN |  |  |  |  |  |  |  |  |  |  |  |  |  |  |  |  |  |  |  | **2** |
| pilO |  |  |  |  |  |  |  |  |  |  |  |  |  |  |  |  |  |  |  | **2** |
| pilP |  |  |  |  |  |  |  |  |  |  |  |  |  |  |  |  |  |  |  | **2** |
| pilQ |  |  |  |  |  |  |  |  |  |  |  |  |  |  |  |  |  |  |  | **2** |
| pilR |  |  |  |  |  |  |  |  |  |  |  |  |  |  |  |  |  |  |  | **2** |
| pilS |  |  |  |  |  |  |  |  |  |  |  |  |  |  |  |  |  |  |  | **2** |
| pilT |  |  |  |  |  |  |  |  |  |  |  |  |  |  |  |  |  |  |  | **2** |
| pilU |  |  |  |  |  |  |  |  |  |  |  |  |  |  |  |  |  |  |  | **2** |
| pilV |  |  |  |  |  |  |  |  |  |  |  |  |  |  |  |  |  |  |  | **2** |
| pilW |  |  |  |  |  |  |  |  |  |  |  |  |  |  |  |  |  |  |  | **2** |
| pilX |  |  |  |  |  |  |  |  |  |  |  |  |  |  |  |  |  |  |  | **2** |
| pilY1 |  |  |  |  |  |  |  |  |  |  |  |  |  |  |  |  |  |  |  | **2** |
| plc1 |  |  |  |  |  |  |  |  |  |  |  |  |  |  |  |  |  |  |  | **2** |
| plc2 |  |  |  |  |  |  |  |  |  |  |  |  |  |  |  |  |  |  |  | **2** |
| plcD |  |  |  |  |  |  |  |  |  |  |  |  |  |  |  |  |  |  |  | **2** |
| rcsA |  |  |  |  |  |  |  |  |  |  |  |  |  |  |  |  |  |  |  | **6** |
| rcsB |  |  |  |  |  |  |  |  |  |  |  |  |  |  |  |  |  |  |  | **8** |
| rfbA |  |  |  |  |  |  |  |  |  |  |  |  |  |  |  |  |  |  |  | **6** |
| rfbB |  |  |  |  |  |  |  |  |  |  |  |  |  |  |  |  |  |  |  | **4** |
| rfbD |  |  |  |  |  |  |  |  |  |  |  |  |  |  |  |  |  |  |  | **5** |
| rfbK1 |  |  |  |  |  |  |  |  |  |  |  |  |  |  |  |  |  |  |  | **4** |
| rpoS |  |  |  |  |  |  |  |  |  |  |  |  |  |  |  |  |  |  |  | **1** |
| sat |  |  |  |  |  |  |  |  |  |  |  |  |  |  |  |  |  |  |  | **2** |
| sciN/tssJ |  |  |  |  |  |  |  |  |  |  |  |  |  |  |  |  |  |  |  | **8** |
| senB |  |  |  |  |  |  |  |  |  |  |  |  |  |  |  |  |  |  |  | **8** |
| sfaC |  |  |  |  |  |  |  |  |  |  |  |  |  |  |  |  |  |  |  | **2** |
| sfaD |  |  |  |  |  |  |  |  |  |  |  |  |  |  |  |  |  |  |  | **1** |
| sfaE |  |  |  |  |  |  |  |  |  |  |  |  |  |  |  |  |  |  |  | **1** |
| sfaF |  |  |  |  |  |  |  |  |  |  |  |  |  |  |  |  |  |  |  | **1** |
| sfaG |  |  |  |  |  |  |  |  |  |  |  |  |  |  |  |  |  |  |  | **1** |
| sfaH |  |  |  |  |  |  |  |  |  |  |  |  |  |  |  |  |  |  |  | **1** |
| sfaS |  |  |  |  |  |  |  |  |  |  |  |  |  |  |  |  |  |  |  | **1** |
| sfaX |  |  |  |  |  |  |  |  |  |  |  |  |  |  |  |  |  |  |  | **1** |
| sfaY |  |  |  |  |  |  |  |  |  |  |  |  |  |  |  |  |  |  |  | **2** |
| sciA |  |  |  |  |  |  |  |  |  |  |  |  |  |  |  |  |  |  |  | **1** |
| sicP |  |  |  |  |  |  |  |  |  |  |  |  |  |  |  |  |  |  |  | **1** |
| sifA |  |  |  |  |  |  |  |  |  |  |  |  |  |  |  |  |  |  |  | **1** |
| sifB |  |  |  |  |  |  |  |  |  |  |  |  |  |  |  |  |  |  |  | **1** |
| sinH |  |  |  |  |  |  |  |  |  |  |  |  |  |  |  |  |  |  |  | **1** |
| sipA/sspA |  |  |  |  |  |  |  |  |  |  |  |  |  |  |  |  |  |  |  | **1** |
| sipB/sspB |  |  |  |  |  |  |  |  |  |  |  |  |  |  |  |  |  |  |  | **1** |
| sipC/sspC |  |  |  |  |  |  |  |  |  |  |  |  |  |  |  |  |  |  |  | **1** |
| sipD |  |  |  |  |  |  |  |  |  |  |  |  |  |  |  |  |  |  |  | **1** |
| Slrp |  |  |  |  |  |  |  |  |  |  |  |  |  |  |  |  |  |  |  | **1** |
| sopA |  |  |  |  |  |  |  |  |  |  |  |  |  |  |  |  |  |  |  | **1** |
| sopB/sigD |  |  |  |  |  |  |  |  |  |  |  |  |  |  |  |  |  |  |  | **1** |
| sopD |  |  |  |  |  |  |  |  |  |  |  |  |  |  |  |  |  |  |  | **1** |
| sopE2 |  |  |  |  |  |  |  |  |  |  |  |  |  |  |  |  |  |  |  | **1** |
| spaO/sctQ |  |  |  |  |  |  |  |  |  |  |  |  |  |  |  |  |  |  |  | **1** |
| spaP |  |  |  |  |  |  |  |  |  |  |  |  |  |  |  |  |  |  |  | **1** |
| spaQ |  |  |  |  |  |  |  |  |  |  |  |  |  |  |  |  |  |  |  | **1** |
| spaR |  |  |  |  |  |  |  |  |  |  |  |  |  |  |  |  |  |  |  | **1** |
| spaS |  |  |  |  |  |  |  |  |  |  |  |  |  |  |  |  |  |  |  | **1** |
| spiC/ssaB |  |  |  |  |  |  |  |  |  |  |  |  |  |  |  |  |  |  |  | **1** |
| sprB |  |  |  |  |  |  |  |  |  |  |  |  |  |  |  |  |  |  |  | **1** |
| sptP |  |  |  |  |  |  |  |  |  |  |  |  |  |  |  |  |  |  |  | **1** |
| ssaC |  |  |  |  |  |  |  |  |  |  |  |  |  |  |  |  |  |  |  | **1** |
| ssaD |  |  |  |  |  |  |  |  |  |  |  |  |  |  |  |  |  |  |  | **1** |
| ssaE |  |  |  |  |  |  |  |  |  |  |  |  |  |  |  |  |  |  |  | **1** |
| ssaG |  |  |  |  |  |  |  |  |  |  |  |  |  |  |  |  |  |  |  | **1** |
| ssaH |  |  |  |  |  |  |  |  |  |  |  |  |  |  |  |  |  |  |  | **1** |
| ssaI |  |  |  |  |  |  |  |  |  |  |  |  |  |  |  |  |  |  |  | **1** |
| ssaJ |  |  |  |  |  |  |  |  |  |  |  |  |  |  |  |  |  |  |  | **1** |
| ssaK |  |  |  |  |  |  |  |  |  |  |  |  |  |  |  |  |  |  |  | **1** |
| ssaL |  |  |  |  |  |  |  |  |  |  |  |  |  |  |  |  |  |  |  | **1** |
| ssaM |  |  |  |  |  |  |  |  |  |  |  |  |  |  |  |  |  |  |  | **1** |
| ssaN |  |  |  |  |  |  |  |  |  |  |  |  |  |  |  |  |  |  |  | **1** |
| ssaO |  |  |  |  |  |  |  |  |  |  |  |  |  |  |  |  |  |  |  | **1** |
| ssaP |  |  |  |  |  |  |  |  |  |  |  |  |  |  |  |  |  |  |  | **1** |
| ssaQ |  |  |  |  |  |  |  |  |  |  |  |  |  |  |  |  |  |  |  | **1** |
| ssaR |  |  |  |  |  |  |  |  |  |  |  |  |  |  |  |  |  |  |  | **1** |
| ssaS |  |  |  |  |  |  |  |  |  |  |  |  |  |  |  |  |  |  |  | **1** |
| ssaT |  |  |  |  |  |  |  |  |  |  |  |  |  |  |  |  |  |  |  | **1** |
| ssaU |  |  |  |  |  |  |  |  |  |  |  |  |  |  |  |  |  |  |  | **1** |
| ssaV |  |  |  |  |  |  |  |  |  |  |  |  |  |  |  |  |  |  |  | **1** |
| ssaX |  |  |  |  |  |  |  |  |  |  |  |  |  |  |  |  |  |  |  | **1** |
| sscA |  |  |  |  |  |  |  |  |  |  |  |  |  |  |  |  |  |  |  | **1** |
| sscB |  |  |  |  |  |  |  |  |  |  |  |  |  |  |  |  |  |  |  | **1** |
| sseA |  |  |  |  |  |  |  |  |  |  |  |  |  |  |  |  |  |  |  | **1** |
| sseB |  |  |  |  |  |  |  |  |  |  |  |  |  |  |  |  |  |  |  | **1** |
| sseC |  |  |  |  |  |  |  |  |  |  |  |  |  |  |  |  |  |  |  | **1** |
| sseD |  |  |  |  |  |  |  |  |  |  |  |  |  |  |  |  |  |  |  | **1** |
| sseE |  |  |  |  |  |  |  |  |  |  |  |  |  |  |  |  |  |  |  | **1** |
| sseF |  |  |  |  |  |  |  |  |  |  |  |  |  |  |  |  |  |  |  | **1** |
| sseG |  |  |  |  |  |  |  |  |  |  |  |  |  |  |  |  |  |  |  | **1** |
| sseJ |  |  |  |  |  |  |  |  |  |  |  |  |  |  |  |  |  |  |  | **1** |
| sseKI |  |  |  |  |  |  |  |  |  |  |  |  |  |  |  |  |  |  |  | **1** |
| sseL |  |  |  |  |  |  |  |  |  |  |  |  |  |  |  |  |  |  |  | **1** |
| sspH2 |  |  |  |  |  |  |  |  |  |  |  |  |  |  |  |  |  |  |  | **1** |
| ssrA |  |  |  |  |  |  |  |  |  |  |  |  |  |  |  |  |  |  |  | **1** |
| ssrB |  |  |  |  |  |  |  |  |  |  |  |  |  |  |  |  |  |  |  | **1** |
| steB |  |  |  |  |  |  |  |  |  |  |  |  |  |  |  |  |  |  |  | **1** |
| steC |  |  |  |  |  |  |  |  |  |  |  |  |  |  |  |  |  |  |  | **1** |
| STM0266 |  |  |  |  |  |  |  |  |  |  |  |  |  |  |  |  |  |  |  | **1** |
| STM0267 |  |  |  |  |  |  |  |  |  |  |  |  |  |  |  |  |  |  |  | **1** |
| STM0268 |  |  |  |  |  |  |  |  |  |  |  |  |  |  |  |  |  |  |  | **1** |
| STM0269 |  |  |  |  |  |  |  |  |  |  |  |  |  |  |  |  |  |  |  | **1** |
| STM0270 |  |  |  |  |  |  |  |  |  |  |  |  |  |  |  |  |  |  |  | **1** |
| STM0271 |  |  |  |  |  |  |  |  |  |  |  |  |  |  |  |  |  |  |  | **1** |
| STM0272 |  |  |  |  |  |  |  |  |  |  |  |  |  |  |  |  |  |  |  | **1** |
| STM0273 |  |  |  |  |  |  |  |  |  |  |  |  |  |  |  |  |  |  |  | **1** |
| STM0274 |  |  |  |  |  |  |  |  |  |  |  |  |  |  |  |  |  |  |  | **1** |
| STM0275 |  |  |  |  |  |  |  |  |  |  |  |  |  |  |  |  |  |  |  | **1** |
| STM0276 |  |  |  |  |  |  |  |  |  |  |  |  |  |  |  |  |  |  |  | **1** |
| STM0278 |  |  |  |  |  |  |  |  |  |  |  |  |  |  |  |  |  |  |  | **1** |
| STM0279 |  |  |  |  |  |  |  |  |  |  |  |  |  |  |  |  |  |  |  | **1** |
| STM0280 |  |  |  |  |  |  |  |  |  |  |  |  |  |  |  |  |  |  |  | **1** |
| STM0281 |  |  |  |  |  |  |  |  |  |  |  |  |  |  |  |  |  |  |  | **1** |
| STM0282 |  |  |  |  |  |  |  |  |  |  |  |  |  |  |  |  |  |  |  | **1** |
| STM0284 |  |  |  |  |  |  |  |  |  |  |  |  |  |  |  |  |  |  |  | **1** |
| STM0285 |  |  |  |  |  |  |  |  |  |  |  |  |  |  |  |  |  |  |  | **1** |
| STM0286 |  |  |  |  |  |  |  |  |  |  |  |  |  |  |  |  |  |  |  | **1** |
| STM0287 |  |  |  |  |  |  |  |  |  |  |  |  |  |  |  |  |  |  |  | **1** |
| sspC |  |  |  |  |  |  |  |  |  |  |  |  |  |  |  |  |  |  |  | **1** |
| tae4 |  |  |  |  |  |  |  |  |  |  |  |  |  |  |  |  |  |  |  | **1** |
| tldeI |  |  |  |  |  |  |  |  |  |  |  |  |  |  |  |  |  |  |  | **1** |
| tcpC |  |  |  |  |  |  |  |  |  |  |  |  |  |  |  |  |  |  |  | **2** |
| tsaP |  |  |  |  |  |  |  |  |  |  |  |  |  |  |  |  |  |  |  | **2** |
| tsh |  |  |  |  |  |  |  |  |  |  |  |  |  |  |  |  |  |  |  | **2** |
| tssA |  |  |  |  |  |  |  |  |  |  |  |  |  |  |  |  |  |  |  | **6** |
| tssB |  |  |  |  |  |  |  |  |  |  |  |  |  |  |  |  |  |  |  | **4** |
| tssC |  |  |  |  |  |  |  |  |  |  |  |  |  |  |  |  |  |  |  | **4** |
| tssF |  |  |  |  |  |  |  |  |  |  |  |  |  |  |  |  |  |  |  | **10** |
| tssG |  |  |  |  |  |  |  |  |  |  |  |  |  |  |  |  |  |  |  | **11** |
| tssJ |  |  |  |  |  |  |  |  |  |  |  |  |  |  |  |  |  |  |  | **4** |
| tssK |  |  |  |  |  |  |  |  |  |  |  |  |  |  |  |  |  |  |  | **4** |
| tssL |  |  |  |  |  |  |  |  |  |  |  |  |  |  |  |  |  |  |  | **4** |
| tssM |  |  |  |  |  |  |  |  |  |  |  |  |  |  |  |  |  |  |  | **5** |
| ugd |  |  |  |  |  |  |  |  |  |  |  |  |  |  |  |  |  |  |  | **9** |
| vasE/tssK |  |  |  |  |  |  |  |  |  |  |  |  |  |  |  |  |  |  |  | **7** |
| vgrG/tssI |  |  |  |  |  |  |  |  |  |  |  |  |  |  |  |  |  |  |  | **1** |
| vipA/tssB |  |  |  |  |  |  |  |  |  |  |  |  |  |  |  |  |  |  |  | **7** |
| vipB/tssC |  |  |  |  |  |  |  |  |  |  |  |  |  |  |  |  |  |  |  | **7** |
| yagV/ecpE |  |  |  |  |  |  |  |  |  |  |  |  |  |  |  |  |  |  |  | **9** |
| yagW/ecpD |  |  |  |  |  |  |  |  |  |  |  |  |  |  |  |  |  |  |  | **9** |
| yagX/ecpC |  |  |  |  |  |  |  |  |  |  |  |  |  |  |  |  |  |  |  | **9** |
| yagY/ecpB |  |  |  |  |  |  |  |  |  |  |  |  |  |  |  |  |  |  |  | **9** |
| yagZ/ecpA |  |  |  |  |  |  |  |  |  |  |  |  |  |  |  |  |  |  |  | **9** |
| ybtA |  |  |  |  |  |  |  |  |  |  |  |  |  |  |  |  |  |  |  | **13** |
| ybtE |  |  |  |  |  |  |  |  |  |  |  |  |  |  |  |  |  |  |  | **13** |
| ybtP |  |  |  |  |  |  |  |  |  |  |  |  |  |  |  |  |  |  |  | **13** |
| ybtQ |  |  |  |  |  |  |  |  |  |  |  |  |  |  |  |  |  |  |  | **13** |
| ybtS |  |  |  |  |  |  |  |  |  |  |  |  |  |  |  |  |  |  |  | **13** |
| ybtT |  |  |  |  |  |  |  |  |  |  |  |  |  |  |  |  |  |  |  | **13** |
| ybtU |  |  |  |  |  |  |  |  |  |  |  |  |  |  |  |  |  |  |  | **13** |
| ybtX |  |  |  |  |  |  |  |  |  |  |  |  |  |  |  |  |  |  |  | **13** |
| ykgK/ecpR |  |  |  |  |  |  |  |  |  |  |  |  |  |  |  |  |  |  |  | **7** |
| **Total** | **71** | **112** | **83** | **86** | **114** | **74** | **158** | **156** | **98** | **53** | **72** | **70** | **52** | **68** | **56** | **20** |  | **19** | **145** | ***** |

This table summarizes the virulence factors (VFs) identified in each of isolates. Each column (BC04–BC38) represents a unique bacterial isolate, while each row lists a specific VF gene. The presence of a virulence gene in an isolate is indicated by a gold–shaded cell. The rightmost column shows the total number of isolates in which each VF was detected.
