## Additional File 4 for "Phenotypic and Genotypic Profile of Enterobacteriaceae Isolated at a Teaching Hospital in Ghana"

**Additional File 4**: Isolates assembly quality

| **Sequence quality after assembly** | | | | | | | | | | | | | | | | | | | | |
| --- | --- | --- | --- | --- | --- | --- | --- | --- | --- | --- | --- | --- | --- | --- | --- | --- | --- | --- | --- | --- |
| **Assembly** | **BC04** | **BC06** | **BC15** | **BC16** | **BC18** | **BC20** | **BC26** | **BC35** | **BC37** | **BC38** | **BC54** | **BC64** | **BC71** | **BC73** | **BC75** | **BC76** | **BC77** | **BC81** | **BC82** | **BC94** |
| Number of contigs (>= 0 bp) | 283 | 1518 | 195 | 628 | 145 | 170 | 257 | 2594 | 471 | 296 | 668 | 356 | 664 | 161 | 768 | 2472 | 1748 | 339 | 324 | 16339 |
| Number of contigs (>= 1000 bp) | 129 | 847 | 63 | 278 | 79 | 50 | 94 | 411 | 99 | 29 | 51 | 73 | 216 | 74 | 257 | 1131 | 151 | 179 | 177 | 1976 |
| Number of contigs (>= 5000 bp) | 87 | 254 | 42 | 166 | 49 | 38 | 69 | 44 | 58 | 20 | 32 | 51 | 136 | 55 | 72 | 472 | 48 | 136 | 137 | 88 |
| Number of contigs (>= 10000 bp) | 76 | 109 | 37 | 132 | 45 | 35 | 59 | 35 | 49 | 19 | 30 | 43 | 102 | 50 | 55 | 261 | 41 | 117 | 115 | 39 |
| Number of contigs (>= 25000 bp) | 52 | 35 | 30 | 90 | 30 | 29 | 36 | 29 | 40 | 19 | 27 | 35 | 76 | 43 | 42 | 91 | 34 | 80 | 82 | 10 |
| Number of contigs (>= 50000 bp) | 31 | 27 | 20 | 59 | 22 | 24 | 29 | 23 | 32 | 15 | 21 | 27 | 55 | 32 | 34 | 37 | 29 | 49 | 48 | 2 |
| Total length (>= 0 bp) | 5103058 | 13154647 | 5335284 | 10630867 | 5188023 | 5400603 | 5377131 | 6839506 | 6199187 | 4823556 | 5888835 | 5496234 | 11303389 | 4904623 | 6438359 | 11186029 | 7738258 | 8945604 | 8938609 | 10099054 |
| Total length (>= 1000 bp) | 5046762 | 12803908 | 5288219 | 10505378 | 5161020 | 5360520 | 5314747 | 5672342 | 6042101 | 4717871 | 5618193 | 5382210 | 11141892 | 4872677 | 6176086 | 10515709 | 6908241 | 8892308 | 8890059 | 4503598 |
| Total length (>= 5000 bp) | 4940979 | 11368443 | 5240797 | 10255732 | 5089522 | 5326460 | 5254567 | 5128309 | 5936121 | 4697549 | 5577593 | 5322737 | 10930259 | 4832616 | 5835795 | 8854628 | 6751073 | 8789351 | 8790719 | 1179077 |
| Total length (>= 10000 bp) | 4860085 | 10354933 | 5203278 | 10016020 | 5062588 | 5304013 | 5188728 | 5069372 | 5876741 | 4688856 | 5562444 | 5258483 | 10684393 | 4794584 | 5726353 | 7389109 | 6701613 | 8656520 | 8632103 | 850204 |
| Total length (>= 25000 bp) | 4461794 | 9269066 | 5100317 | 9320339 | 4827099 | 5218938 | 4821292 | 4958119 | 5715463 | 4688856 | 5495572 | 5140454 | 10270506 | 4696425 | 5497886 | 4831084 | 6607848 | 8023984 | 8070584 | 406289 |
| Total length (>= 50000 bp) | 3684974 | 8993806 | 4724209 | 8243714 | 4534368 | 5051621 | 4564714 | 4724724 | 5424050 | 4546090 | 5281996 | 4873866 | 9512881 | 4323358 | 5188641 | 2957155 | 6408681 | 6888733 | 6838747 | 108324 |
| Number of contigs | 158 | 1160 | 86 | 351 | 95 | 64 | 121 | 1485 | 144 | 45 | 183 | 101 | 269 | 88 | 477 | 1649 | 818 | 196 | 202 | 5051 |
| Largest contig | 251190 | 1647026 | 719109 | 363775 | 499103 | 797452 | 346426 | 466445 | 490738 | 736846 | 1213408 | 758250 | 808235 | 358071 | 416076 | 172861 | 690920 | 487665 | 512829 | 54888 |
| Total length | 5066541 | 13022113 | 5302022 | 10556077 | 5172106 | 5369925 | 5333584 | 6400268 | 6070867 | 4727661 | 5696478 | 5399715 | 11178202 | 4881803 | 6326706 | 10854432 | 7347600 | 8903532 | 8906737 | 6617473 |
| GC (%) | 50.55 | 45.45 | 50.49 | 54.01 | 50.64 | 57.42 | 50.73 | 50.52 | 56.39 | 52.24 | 57.01 | 50.31 | 55.64 | 50.57 | 56.16 | 55.66 | 64.05 | 45.35 | 45.35 | 56.78 |
| N50 | 113358 | 295243 | 287844 | 138630 | 229264 | 385507 | 174925 | 241691 | 203945 | 389688 | 308031 | 263083 | 182186 | 135086 | 159470 | 20076 | 307628 | 149031 | 144232 | 1519 |
| N90 | 21041 | 3808 | 49805 | 22511 | 41651 | 66773 | 29592 | 886 | 47892 | 122540 | 79276 | 51252 | 31211 | 42069 | 11721 | 2774 | 16169 | 25047 | 25788 | 614 |
| auN | 110380.2 | 434486.5 | 372136.1 | 149629 | 270082.2 | 384752.5 | 179704.8 | 203855.2 | 213404.4 | 420438.4 | 484005 | 291123.7 | 273431.7 | 156015.6 | 179394.9 | 35676.5 | 310705.3 | 166860.2 | 179336.4 | 5348.8 |
| L50 | 16 | 12 | 5 | 25 | 7 | 5 | 11 | 10 | 11 | 5 | 5 | 7 | 15 | 12 | 12 | 118 | 8 | 19 | 18 | 998 |
| L90 | 57 | 335 | 21 | 98 | 25 | 21 | 36 | 505 | 33 | 12 | 19 | 27 | 69 | 34 | 53 | 720 | 35 | 80 | 80 | 3849 |
| # Ns per 100 kbp | 0 | 0 | 0 | 0 | 0 | 0 | 0 | 0 | 0 | 0 | 0 | 0 | 0 | 0 | 0 | 0 | 0 | 0 | 0 | 0 |
